# Predictors of Adolescent Engagement and Outcomes – a cross-sectional study using the Togetherall (formerly Big White Wall) digital mental health platform

**DOI:** 10.1101/2021.08.20.21262337

**Authors:** Nushka Marinova, Tim Rogers, Angus MacBeth

**Author notes:** Corresponding Author: Dr Angus MacBeth, School of Health in Social Science, The University of Edinburgh, Rm 2.11, Doorway 6, Medical Quad, Teviot Place, Edinburgh, EH8 9AG.

## Abstract

Online mental health platforms can improve access to, and use of, mental health support for young people who may find it difficult to engage with face-to-face delivery.

We modelled engagement and change in anxiety and depression symptoms in adolescent users of the Togetherall (formerly “Big White Wall”) anonymous digital mental health peer-support platform.

A cross-sectional study assessed online activity data from members of Togetherall in UK adolescents referred from mental health services (N=606). Baseline demographics, depression, anxiety, and usage statistics were assessed. Symptom levels among participants who chose to take validated anxiety and depression measures were measured. And participant characteristics were used to predict engagement.

Mean number of logins for adolescent members was higher for older adolescents, and for a longer duration than younger adolescents. Mean number of logins and usage time was higher in female adolescents than males. For the total sample, 47.9% of users accessed more than one course, and 27% accessed at least one self-help resource. Gender and age predicted number of joined courses. Greater accessed self-help materials predicted reduced anxiety symptoms. Members’ mean baseline symptom levels were: GAD-7 between 13.63 and 14.79; PHQ-9 between 16.8 and 18.58.

Data were derived from a naturalistic design and modelling of multiple symptom scores should be interpreted with caution.

Findings show that adolescents readily engage with an anonymous online platform for common mental disorder, with scope for tailored pathways for different symptom profiles. Members benefit from engagement with Togetherall materials and courses.

---

Mental health problems are a leading cause of youth disability (^1^) with global estimates of 6.5% for anxiety, 2.5% for depression and 13.4% for mental health problems in young people (^2^). Untreated mental disorders negatively affect interpersonal relationships, academic attainment, increasing the risk of adult psychological problems (^3,4^). Timely, effective early mental health interventions may improve treatment response and long-term outcomes (^5,6^). However, over half of young people do not access or receive psychological treatment (^7,8,9^). In the UK, only 25% of those referred to specialist Child and Adolescent Mental Health Services (CAMHS) receive treatment, alongside long waiting times (^10^). Despite initiatives demand continues to outstrip capacity (^11,12^). The treatment gap may also reflect adolescents’ preferences for self-management and stigma-related concerns, which inadvertently create barriers to accessing mental health treatment (^13,14^).

E-mental health, encompassing psychological approaches digitally delivered via internet and mobile devices, may promote better mental health in young people who may otherwise not access support. Advantages include wider population reach; flexible and convenient access; privacy and anonymity (reducing stigma); scalability and lower cost delivery (^15–17^). Evidence indicates pervasive use of digital mental health services by young people, (^18,19^) with high acceptability, satisfaction (^20,21^) and treatment efficacy for youth digital interventions targeting anxiety and depression (^16,22–25^); with stronger support for online Cognitive Behavioural Therapies (CBT) relative to other digital interventions (^26,27,28^). However, there is significant heterogeneity between studies (^29^). Transdiagnostic approaches for young people may be as efficacious as disorder specific interventions in e-mental health research (^25^), with large effect sizes compared to no intervention (^26,29^), and in providing treatment for, and prevention of common mental health problems (^29,33,34^). E-mental health interventions incorporating guidance also have greater effectiveness and lower drop-out rates in young people than non-guided interventions (^25,29,32^).

Attrition is also a widely reported issue in e-mental health (^16,33^). Therefore understanding engagement with digital content may improve intervention adherence (^34^). Better engagement with digital interventions predicted improved outcomes for common mental health problems in both young people (^16,35–37^) and adults (^38,39^). However, there is heterogeneity in how engagement is operationalized (e.g. usage time, login counts and number of completed modules (^36,40^). Furthermore, age (^27,41,42^) and female gender (^35,43,44^) have been associated with better treatment response and greater adherence to e-mental health interventions, whereas findings for baseline symptom severity have been more equivocal (^3,7,42,44,45,46^).

One candidate digital mental health approach is Togetherall (formerly Big White Wall, ‘BWW’), a multimodal platform incorporating a community of members providing each other with anonymous, round-the-clock peer support made safe by professionally registered moderators, self-help materials, guided courses, digital art and self-monitoring of psychological symptoms (^47^). Togetherall’s self-help materials and group courses cover a variety of psychological and wellbeing topics informed by evidence-based approaches (^47^); adopting a transdiagnostic approach towards mental health. Tailored care may be particularly valuable for adolescents, given their preference for personalised digital services (^48^). Togetherall’s offers a community of peers providing mental health support to each other, where members create group discussions, anonymous friendship online and join dedicated group fora for different shared courses into which they enrol. Peer support features facilitate young people’s engagement with other web-based interventions (^49^), reduce perceived isolation (^50^); whilst use of peer networking enhances online intervention adherence (^49^) and provides opportunities for social support and connection (^51,52^).

Togetherall is unique within mental health technology solutions as members are kept safe within the community via a combination of basic AI, monitoring of online posts, and through continuous moderation by professionally registered mental health practitioners (“Wall Guides”). Moderators have immediate access to senior clinicians. Togetherall refers members at imminent risk to local crisis or emergency services internationally when needed. It is therefore a safe, low risk environment for young people to support each other’s mental health. Moderation can also be used to address potential overdependency (^53^) and emotional contagion in web-based peer-support networks (^54^), enhancing the quality and safety of e-mental health support services (^52^). Narrative and RCT evidence supports the potential of Togetherall (^50,55^), with evidence for significant increases in recovery, and decreased anxiety/depression symptoms after use (^56^). Greater use of Togetherall (‘BWW’) in the study was associated with larger improvements in anxiety and depression outcomes (^56^).

The current study explored adolescent members’ engagement with the platform and modelled baseline symptoms by age and gender. We investigated anxiety and depression symptom levels in these users of hypothesizing that gender and age would predict usage and symptom outcomes; and females and older participants would display greater use of the platform and subsequent greater reduction in initial symptoms compared to males and younger users. We also hypothesised that higher baseline anxiety and lower baseline depression symptoms would predict greater use of Togetherall and symptom reduction. Greater usage metrics and access of intervention components were expected to predict lower final anxiety and depression symptom scores.

## Methods

### Study Design and Ethics

A cross-sectional design explored use of routinely collected online activity data from Togetherall. Registered members’ agreement with the Togetherall (BWW) “Terms of Use” formed semi-passive consent to use of data for the research. Processing of data for research purposes was anonymized according to applicable data protection laws. A data sharing agreement was agreed and the study approved by University of Edinburgh School of Health in Social Science Ethics Committee.

### Sample

Data were from Togetherall users referred from UK CAMHS (N=1693). Users were excluded if they were >18 years old (N=678). Excluding logins <60 seconds and participants without symptom measures gave a final sample of N=607. Participants were aged between 16 and 18 years (M=17.3, SD=0.71), predominantly female (78.7%) and White British (84.3%). Twenty participants (3.3%) reported “other” or “non-specified” gender”.

#### Procedure

Data were extracted from a Mini SQL database system and comprised anonymized demographic characteristics (gender, age, ethnicity, referrer); login times and duration; questionnaires taken; guided support courses and self-help pages accessed on Togetherall. All measures were anonymised self-reports. Data were based on user log-ins to Togetherall from August 2016 to June 2019. Collected usage metrics were averaged for each participant. Completed anxiety and depression questionnaires on the website were taken as baseline symptoms. For outcomes, scores were considered from first to last completed anxiety and/or depression questionnaire. Number of questionnaires taken per participant was also recorded. All participants’ data were evaluated for engagement. For treatment outcomes, data were included from participants who completed >1 anxiety (N=200) or depression (N=245) questionnaire (Figure 1). The majority of the sample (N=606) did not take a clinical test. Members choose when and whether to take a clinical test depending upon their personal motivation to do so at different points in their individual user journeys.

**Figure 1.**
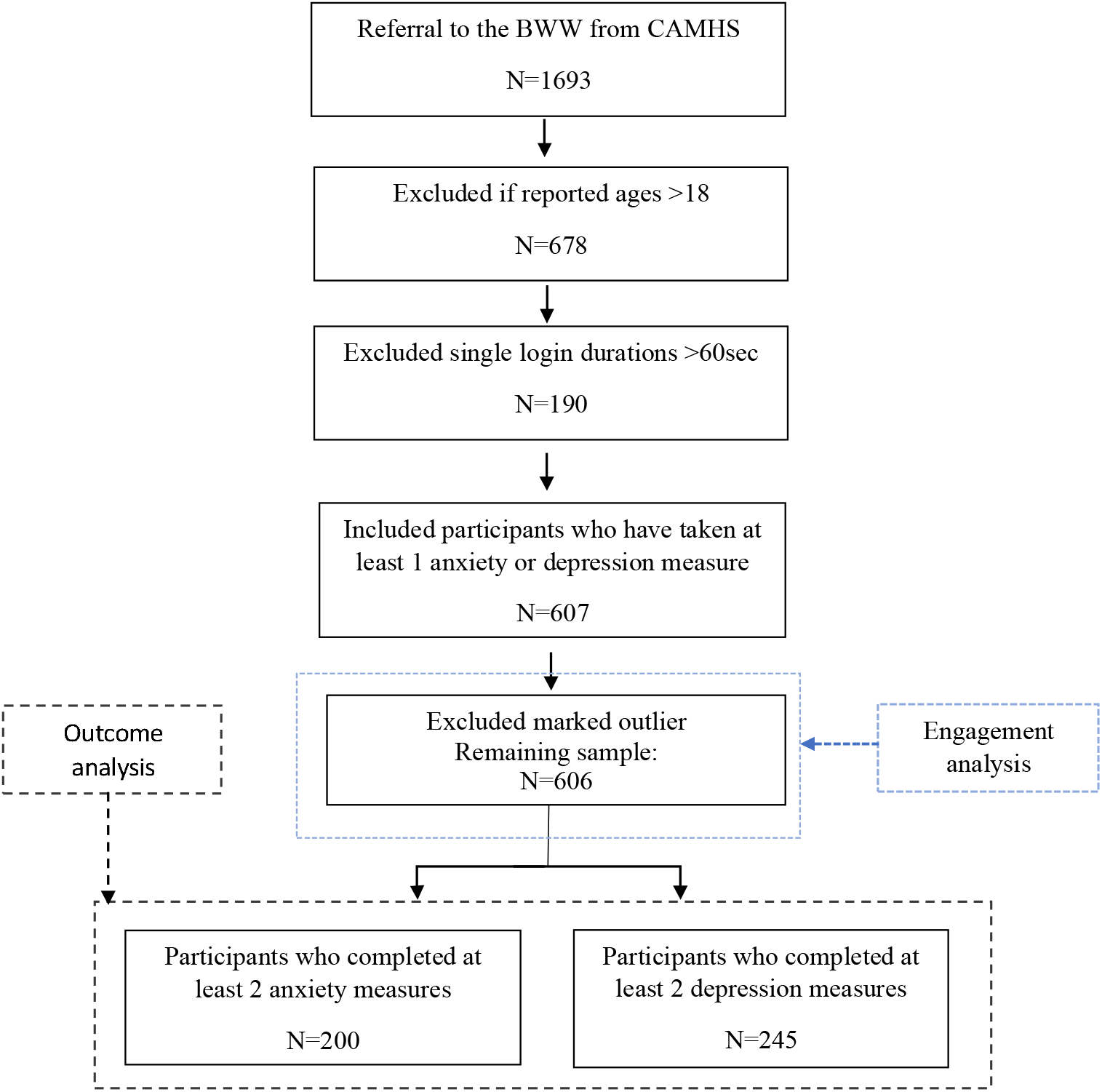
Participant flow.

### Intervention

Togetherall is a multimodal web-based platform for managing psychological distress and improving wellbeing. It offers a 6-month renewable membership and free access for certain user groups (e.g. universities or state territories). Materials and programmes are based on self-management techniques, informed by evidence-based practice including CBT (^47,57,58^). Registered mental health professionals continuously moderate Togetherall forums and offer guidance to users. Togetherall also sends automatic notifications of online activity to members. Service components include guided courses (“Guided Support”), self-help materials (“Useful Stuff”), peer support forum (“The Community”) and platform for creating digital art (“Bricks”). Separate from these services, some organisations also commission online psychotherapy (“Live Therapy”). Togetherall’s “Guided support” offers free structured programmes on mental health and general wellbeing, lasting 2 to 8 weeks. Members can sign up for multiple courses, opt-out and choose when to do activities. Enrolled participants receive weekly course activities, notifications and email prompts and are encouraged to use peer support feature from a dedicated course forum. Self-help materials include psychological and health education and advice on skills development. These materials are organized into 8 categories, including emotional health, life-skills, health and lifestyle. Togetherall further offers voluntary self-monitoring of wellbeing on a large number of validated measures (e.g. depression, anxiety, self-esteem). Members are encouraged to complete routine anxiety and depression measures, at first login (baseline) and throughout completion of Togetherall activities.

### Measures

#### Usage measures

Total usage time was measured by combining the duration of an individual’s BWW logins (minutes). Logins of < 1 minute were excluded and interactions longer than 1 hour were coded 1 hour to limit the contribution of idle periods towards the total count, consistent with other operationalizations of engagement on digital platforms (^59^). Number of logins denoted the total number of times each user accessed Togetherall. Average user time per session (minutes) was defined as an individual’s total usage time divided by number of logins. Number of courses taken was derived by summing all “Guided Support” courses users joined. Number of accessed self-help materials was an additive measure of all opened self-help materials per participant. Zero counts were assigned to users who did not access Togetherall self-help or guided support courses. Time registered (minutes) was calculated as the time difference between first and last access of BWW for each participant.

#### Outcome measures

##### Anxiety and Depression

The primary anxiety outcome was the Generalized Anxiety Disorder Screen (GAD-7) (^60^) which has established general population reliability and validity (^61^), evaluating anxiety over the past 2 weeks. The maximum score is 21, with cut-off points of 5, 10, 15 representing mild, moderate and severe levels of anxiety and a score greater than 10 indicating a diagnosis of generalized anxiety disorder (^60^). Depressive symptoms were measured via the Patient Health Questionnaire, PHQ-9 (^62^), which has good general population reliability and validity (^63^). The maximum score is 27, a score of 10 or greater suggest a diagnosis of major depression and scores of 5, 10, 15 and 20 represent mild, moderate, moderately severe and severe depression symptoms (^62^). Internal consistency of the PHQ-9 and GAD-7 were not calculated due to database format.

### Data Analysis

All analyses were performed in R. For engagement, the response variables of interest were usage time (in minutes), number of logins, number of courses and number of self-help materials accessed. Demographic predictor variables of interest were gender, age, baseline anxiety and depression. The predictor variables gender (Levels: Male, Female, Other) and age (Levels: 16, 17, 18) were categorical and dummy-coded, taking the first level (Gender=Male, Age=16) as reference categories with remaining variables treated as continuous. Total time (in minutes) of registration with Togetherall was entered as a control predictor. Average session duration (minutes) was included as a covariate in the analysis of total usage time to control for individual session duration variability. Multicollinearity diagnostics (^64^) detected collinearity between anxiety and depression but all Variance Inflation Factors were < 2.5 thus the variables were retained as predictors.

There were missing observations for 14% of baseline anxiety (N=86) and 6% of baseline depression (N=37). Multiple imputation was implemented via classification, regression trees (^65^) and the “mice” package (^66^). Visual inspection of distributions of original against imputed data showed good approximation of imputed values to observed values with identical means and standard deviations (Supplemental Material Figure 2). For anxiety outcomes, there were 200 observations with 4 missing baseline values imputed. Depression outcome data consisted of 245 observations and 10 imputed baseline scores.

**Figure 2:**
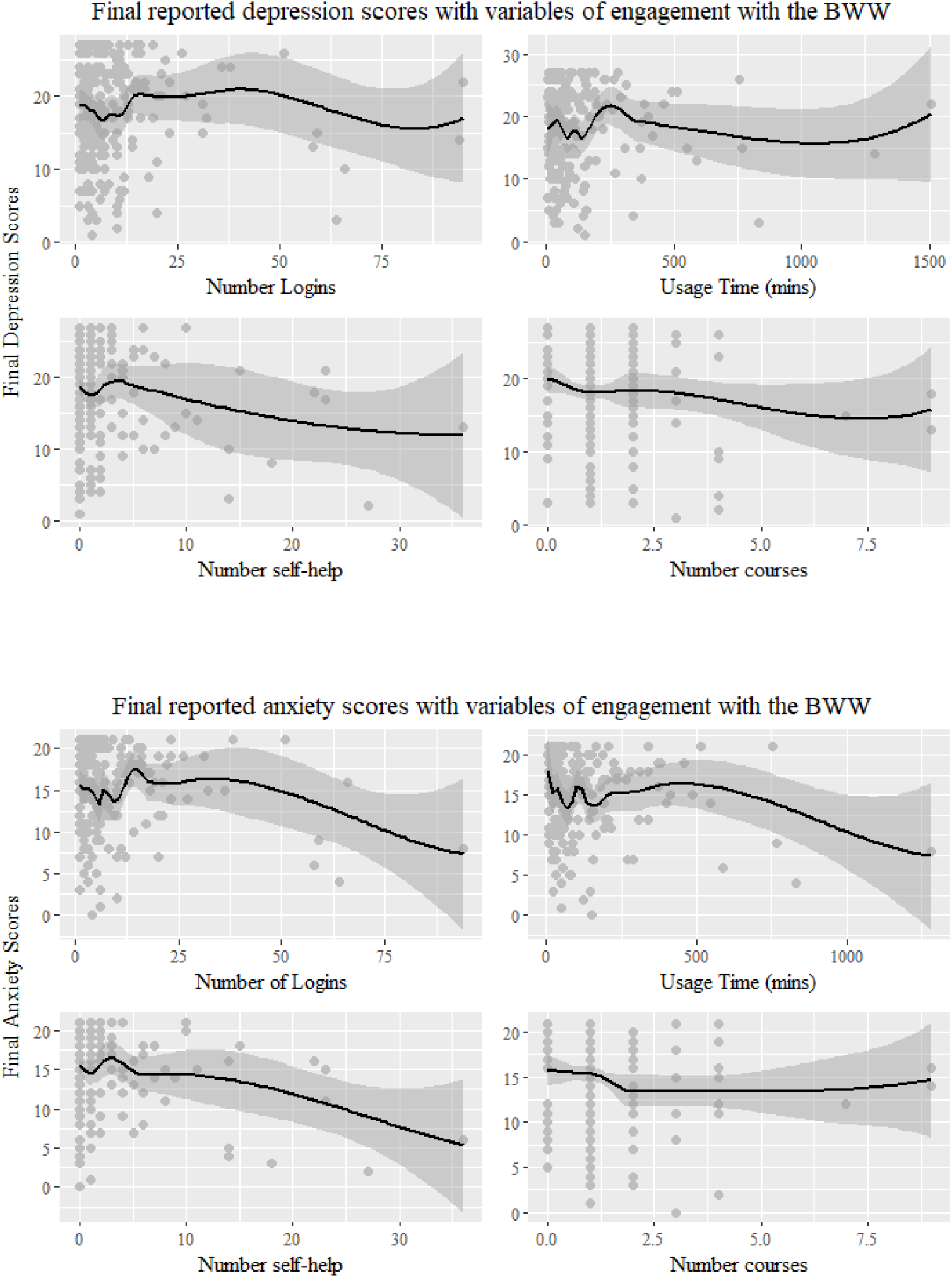
Regression line trends for anxiety and depression.

Due to violations of assumptions for multiple linear regression, predictors of total usage time were modelled using robust regression with bootstrapped coefficients. Generalized linear models with negative binomial distribution were fitted to estimate login count, number of accessed self-help and number of accessed courses. Model fit diagnostics through “DHARMa” (^67^), showing improved fit of the negative binomial models and no significant over-dispersion, compared to Poisson regressions with the same variables. To evaluate symptom change, only participants with ≥2 measures for anxiety or depression were retained for outcome analyses. Last obtained anxiety or depression questionnaire scores from each participant were taken as response variables, including baseline scores as covariates (^68^). Individual diagnostic tests indicated multicollinearity and highly correlated predictors between usage time and number of logins (VIF>10.5). Therefore, number of logins was dropped as an outcome predictor. Stepwise linear regressions with 10-fold cross-validations were performed to select optimal models with the least prediction error using “caret” (^69^).

## Results

### Overview

Visual inspection identified one outlier, which was removed from analyses. Remaining data remained dispersed, consistent with observational studies of real-world engagement with online services (^33,59,70^). To evaluate potential bias in the model estimates, analyses were conducted with influential outliers removed, (Cook’s distance >4x the mean of each fitted model (^71,72^)). Full models are reported, as there were few significant differences in R-squared models and distribution of fitted and residual values (see Supplemental Material Tables 3-5 for analyses including outliers).

Participants with only one log-in represented 33.5% of the total sample. Mean number of logins was 5.11 (SD=8.93), with a mean usage time of 64.22 minutes (SD=123.27). Members who accessed one or more guided courses represented 47.9% of the sample (n=290) and 27% (n=164) accessed more than one self-help page. Baseline scores indicated moderate anxiety (M=14.57, SD=4.58) and depression (M=17.8; SD=5.79). There were no significant differences in baseline anxiety (F=.28(2, 480), p=.78) and depression scores (F=.702(2,480), p=.51) between age groups (Table 1). Tukey HSD post-hoc tests demonstrated members identifying as female or “other” gender had significantly higher baseline depression scores than males (F=4.96 (2, 480), p=.007) (Table 1).

**Table 1.**
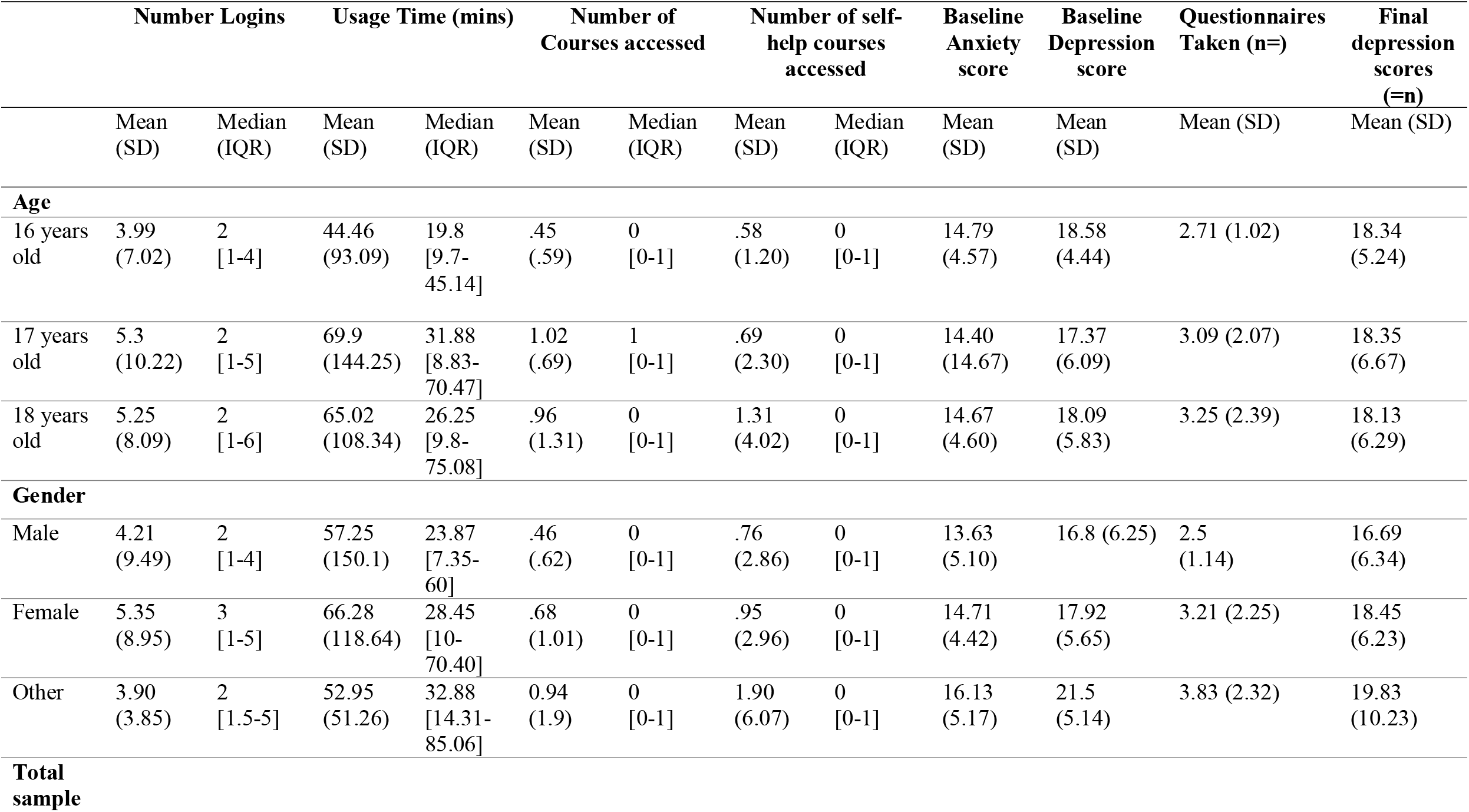
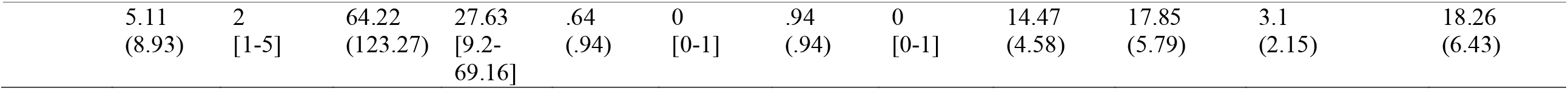
Descriptive statistics of engagement variables and baseline symptoms by age and gender (n=607)

### Predictors of usage time

Multiple linear regression predicting usage time was conducted using age, gender, baseline depression and anxiety as candidate predictor variables. The control variables were total time registered and average session duration (minutes). Visual examination of plotted residuals for log-transformed usage time indicated violations of homoscedasticity and non-linearity. Regression coefficients were bootstrapped with random resampling over 2000 replications (^73^). Original and bootstrapped density estimates with plotted confidence intervals are depicted in Supplemental Material (Supplemental Figure 1). Table 2 displays the coefficients, standard errors and Bootstrap confidence intervals. Results indicated that the control predictor variables of average session duration (b=2.53 (1.914, 3.086), p<.001) and total time of Togetherall access (b<.001 (<.001, 001)) were significantly associated with usage time. No other variables emerged as significant predictors.

**Table 2.**
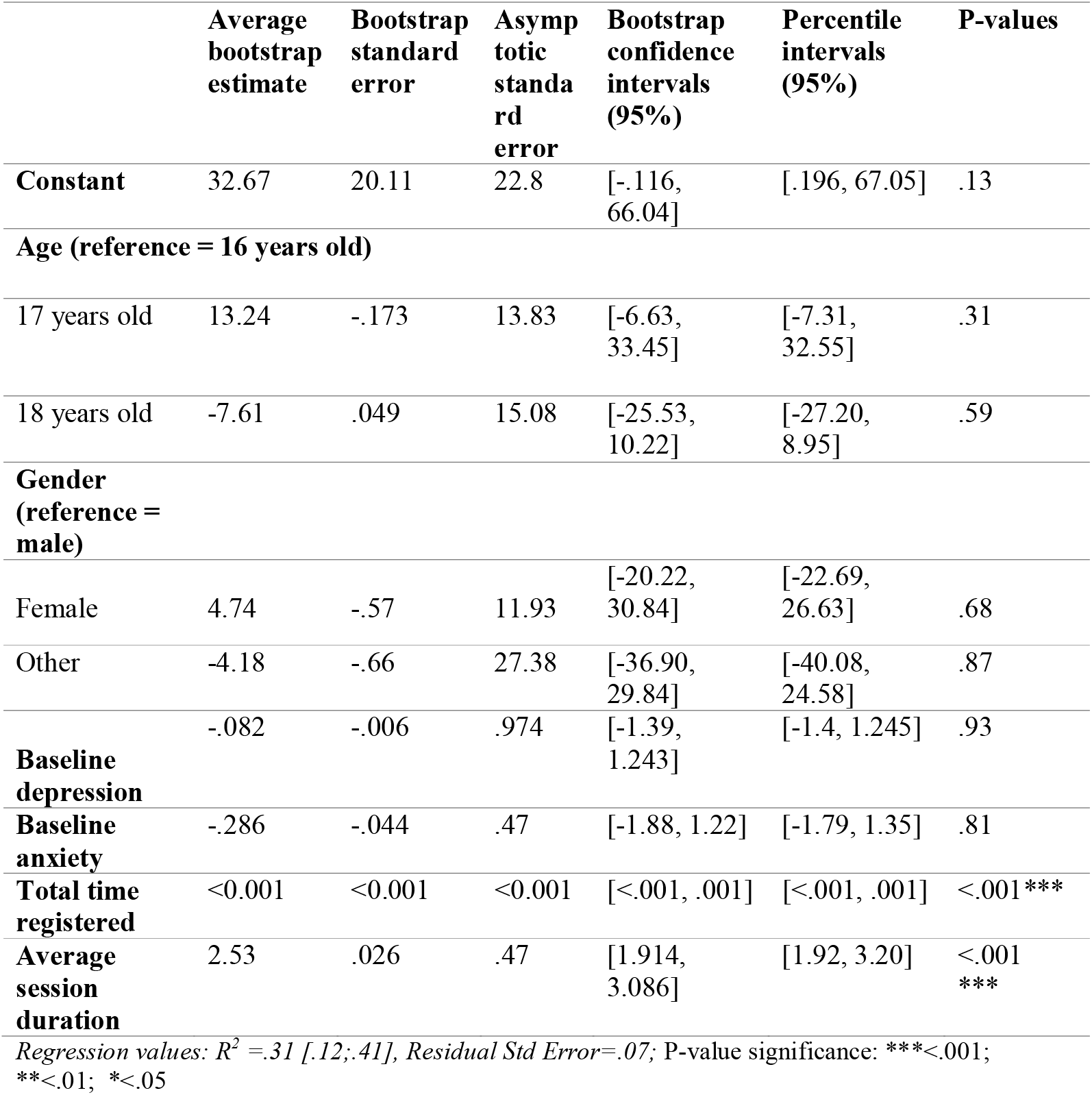
Robust regression coefficients for predicted usage time

**Table 3.**
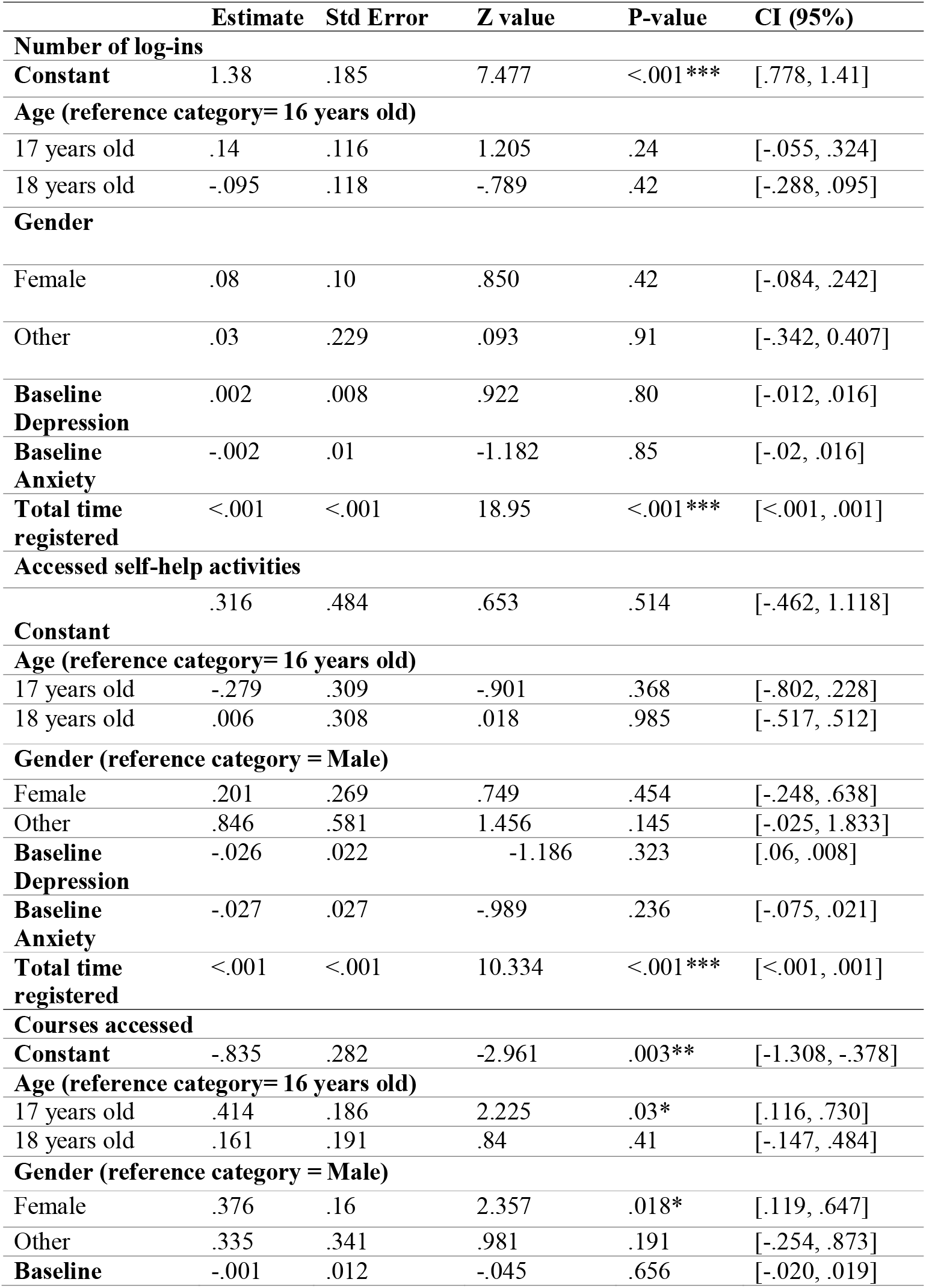
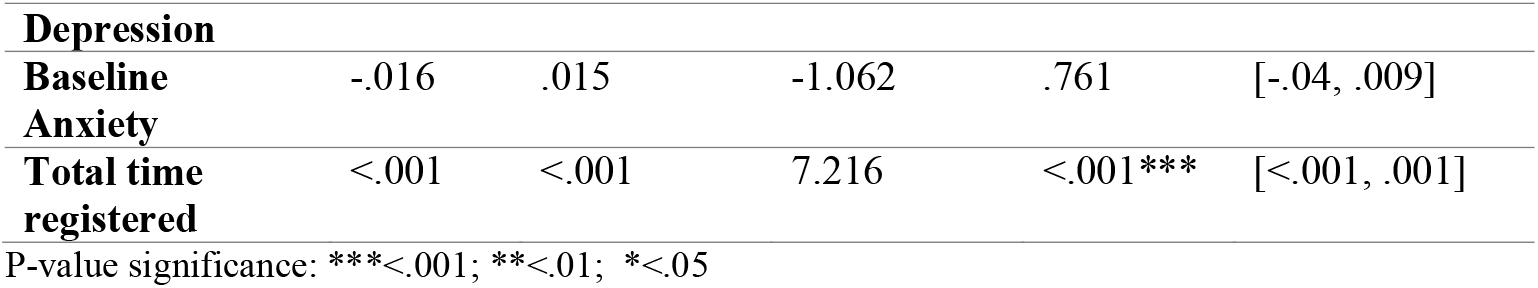
Negative binomial regressions for predicted log-ins, number of self help activities accessed, and number of courses taken (counts)

### Predictors of number of logins

Due to over-dispersion in the initial Poisson regression, a negative binomial model with a log-link function was fitted to predict number of Togetherall logins. Predictors of interest were age, gender, baseline depression and anxiety, with total time registered (minutes) as the control predictor. Non-parametric tests comparing fitted to the residual values, indicated no significant overdispersion in the negative-binomial model (Ratio=1.32, p=.288). Based on the residual deviance test with a chi-squared distribution (critical cut-off for residual deviance, G^2^=656), a negative binomial regression showed better model fit (G^2^=556.2, DF=598, p=.89), compared to a Poisson regression model with the same predictors (G^2^=2995, DF=589, p<.001). No predictors of interest predicted log number of logins (Table 3). The control variable of total time registered was significant, showing that expected logins increased as time spent accessing Togetherall increased (Table 3).

### Predictors of accessed self-help and guided support

Negative binomial models were used to estimate frequency of use of self-help and courses joined, using the same set of predictors as the model for number of logins. Negative binomial modes were selected due to indicated over-dispersion in the initially fitted Poisson regressions. The nonparametric dispersion test of the ratio between residual and fitted values indicated no overdispersion in the negative binomial regressions predicting number of accessed self-help (Ratio=.81, p=.94) and number of accessed courses (Ratio=1.06, p=.184). Goodness of fit metrics, based on the residual deviance test (critical deviance value, G^2^=656), showed appropriate fit for the data from the self-help (G^2^=374.11, DF=598, p=.99), and accessed course models (G^2^=559.9, DF=598, p=.87). The control variable of total time of access to Togetherall was a significant predictor of greater log counts for both self-help and courses activity (Table 3). No other variables significantly predicted accessed self-help. For courses joined, gender and age emerged as significant predictors (Table 3). The log count of courses increased by 0.4 counts for female compared to male Togetherall users, (b=.376 (.119, .647), p=.02) and by 0.4 counts in 17-year-old compared to 16-year-old members (b=.414 (.116, .730), p=.03).

### Outcome analyses

#### Depression

Outcomes were modelled from users who had completed ≥2 depression measures (N=245). Within this subsample, on average, members spent 109 minutes on Togetherall with a mean of 8 logins (Supplemental Table 1). For the whole subsample, 43.7% (107/245) viewed more than one Togetherall self-help page whereas 81.2% (199/245) joined one or more courses. Baseline and final depression measures were analysed, including number of measures taken as a covariate. As for the full sample, baseline scores indicated moderate levels of anxiety and depression (M=14.38, SD=4.47; M=17.88, SD=5.74, respectively)/ There were no significant differences in baseline depression or anxiety scores between the different age and gender groups. Data trends from the regression line indicated a slight decline in final depression scores for members who accessed more self-help and courses (Figure 2).

Stepwise regression with forward selection was applied to identify the best performing model for prediction of final depression scores. Ten-fold cross-validation was used to estimate the average prediction error (RMSE) and the model with best number of significant predictors and lowest RMSE was selected. Number of Logins was dropped before selection due to high correlations with Usage Time. Selection started from a full model of 9 predictors, including age, gender, baseline anxiety and depression, usage time, average session duration, number of accessed courses and self-help and the control predictors number of taken questionnaires and total time being registered. The best performing model included predictors of accessed self-help and courses, baseline depression and anxiety scores, total usage time and average time spent per login (Table 4). Depression, (b=.772 (.684, .861), p<.001), and anxiety scores, (b=.179 (.065;.293), p=.009), at baseline significantly predicted higher final depression scores. Total usage time (b=.004 (.001,.006), p=.02) and average session duration (b=.061(.013;.108), p=.03) significantly predicted increased final depression scores by a mean of 0.004 and 0.06 points respectively (Table 4).

**Table 4:** Stepwise regression coefficients for predictors of final depression (n=245) and anxiety scores (n=200)

|  | **Estimate** | **Std Error** | **T value** | **P-value** | **CI (95 %)** |
| --- | --- | --- | --- | --- | --- |
| **Final Depression score** |  |  |  |  |  |
| **Constant** | .086 | 1.267 | .068 | .946 | [-2.007, 2.178] |
| **Gender** (reference category = male) |  |  |  |  |  |
| Female | 1.316 | .763 | 1.724 | .086 | [-.055, 2.576] |
| Other | .997 | 1.806 | .552 | .581 | [-1.986, 3.98] |
| **Baseline anxiety** | .179 | .069 | 2.598 | .009** | [.065, .293] |
| **Baseline depression** | .772 | .054 | 14.385 | <.001*** | [.684, .861] |
| **Usage time** | .004 | .002 | 2.392 | .02* | [.001, .006] |
| **Average session duration** | .061 | .029 | 2.121 | .03* | [.013, .108] |
| **Number of accessed self-help** | -.109 | .067 | -1.645 | .101 | [-.220, .001] |
| **Number of accessed courses** | -.329 | .246 | -1.336 | .183 | [-.735, .078] |
| **Final Anxiety Score** |  |  |  |  |  |
| **Constant** | 2.986 | .889 | 3.359 | .001** | [1.517, 4.455] |
| **Baseline Anxiety** | .661 | .062 | 10.613 | <.001*** | [.558, .764] |
| **Baseline Depression** | .149 | .045 | 3.295 | .001** | [.074, .223] |
| **Number of accessed self-help** | -.115 | .047 | -2.477 | .014* | [-.192, -.038] |

P-value significance: \*\*\*<.001; \*\*<.01; \*<.05. Model fit for depression: $R^2=.62$ , Std Error=3.97, DF=236; Model fit for anxiety: $R^2=.56$ , Std Error=3.21, DF=196

#### Anxiety

Outcomes for anxiety scores were modelled from users with ≥2 anxiety measures. Mean usage time for Togetherall in this sub-sample was 117 minutes, with a mean frequency of 9 logins (Supplemental Table 2). Within this subsample, 46% (92/200) accessed more than one Togetherall self-help resource and 84% (168/200) joined more than one course. Baseline and final anxiety scores were analyzed with number of taken questionnaires included as covariate. Moderate levels of mean baseline anxiety and depression scores were observed (M=14.64, SD=4.35; M=17.42, SD=6.01, respectively) (Supplemental Table 2). There were no significant differences in baseline depression or anxiety scores between the different age and gender groups. Regression lines indicated a decrease in final anxiety scores associated with higher logins and usage time (Figure 2). However, it appeared biased towards data points with >50 logins and >8 hours on Togetherall. The regression line for self-help materials illustrated a gradual decline in final anxiety scores with greater accessed self-help frequency. No clear patterns emerged for the distribution of anxiety scores against number of courses accessed, with equal distribution of low and high anxiety scores across different number of joined courses.

Stepwise regression with forward selection and 10-fold cross-validation was conducted to select the best performing model with lowest prediction error (RMSE). Number of Logins was highly correlated with Usage time and was omitted from model selection. Selection used the same 9-predictors utilized for the depression analyses. The best performing model included baseline depression and anxiety and number of accessed self-help, all of which significantly predicted changes in final anxiety scores (Table 4). Higher baseline anxiety and depression predicted higher final anxiety scores by 0.7 and 0.15 points respectively). Increasing number of accessed self-help was associated with a .12-point decrease in final anxiety scores, (b=-.115, (-.192, -.038), p=.013).

### Sensitivity analysis: Complete-case analysis

Sensitivity analyses omitting missing data showed gender significantly predicted greater usage time (Supplemental Table 6) and number of logins (Supplemental Table 7); whereas age was no longer a significant predictor of accessed number of courses (Supplemental Table 7). For final depression scores, average session duration did not significantly predict final depression scores while usage time remained a significant predictor (Supplemental Table 8). The models estimating number of accessed self-help and anxiety remained unchanged (Supplemental Tables 7&8).

## Discussion

The current study explored predictors of engagement and outcomes for CAMHS referred users of the Togethreall digital peer-support platform. Results showed that females accessed significantly more guided courses than males, and subsequently reported lower symptoms.. Findings showed adolescents readily engaged with an anonymous online platform to manage common mental disorders. Baseline symptom scores among adolescents using the service were significan, generally in the moderately severe range for depression and anxiety. The Togetherall platform therefore reaches a group of adolescents with significant levels of morbidity, at diagnostic levels of severity, rather than indicative of a simple expression of distress. However, the data suggests that adolescents sign up to the service before their symptoms become clinically severe.

Most members did not complete more than one clinical test in their journey through the service. For he minority of members who did, our hypotheses about baseline anxiety and depression scores did not predict changes in engagement with Togetherall. Accessing more Togetherall self-help materials predicted reduced final anxiety scores, but no other engagement or demographic variables predicted changes in symptom outcomes.

Total usage time and logins were not significantly predicted by participants’ characteristics, suggesting similar overall usage of Togetherall in adolescents regardless of baseline demographics and symptom severity. However, these measures of engagement do not capture the extent to which users meaningfully engage with digital support (^34,36,40^). For engagement with specific Togetherall components, female gender predicted greater use of guided courses, consistent with evidence that female adolescents show greater module completion in web-based psychological interventions (^35^) and better help-seeking intentions towards e-mental health services (^74^,^75^). This may also reflect variance in preferences for different Togetherall features.

Our sample included a narrower age range than comparable studies of age in digital mental health (^27,41,42^), but were similar to existing findings for young people’s interactions with e-mental health (^74,76^). Contrary to our findings for Togetherall component guided courses, we identified no predictors for accessing Togetherall self-help materials. This contrasts with existing evidence for associations between demographic characteristics and intervention adherence in web-based interventions (^36^). The Togetherall self-help feature may thus represent a different modality for e-mental health support. Further, meta-analytic data reports limited influence of patient characteristics on effect sizes of self-help interventions for anxiety and depression in youth (^77^) consistent with our results predicting Togetherall self-help material usage.

Baseline anxiety and depression did not predict engagement with Togetherall components, contrasting with existing research linking greater e-mental health adherence to lower depression and higher pre-treatment anxiety (^46,78^). However, evidence is equivocal, with evidence for associations in contradictory directions (^35,37,42,79^). One possibility is that initial symptom severity may not predict adolescents’ Togethereall usage, or the relationship between symptom severity and engagement may be moderated by other factors. Beyond individual characteristics, amount of guidance (^25,29,80^), on-platform peer support (^49^) and web-design features (^81,82^) may influence digital intervention adherence. Additional digital design variables may therefore be important in predicting engagement and understanding wellbeing improvements related to Togetheral use.

For symptom change, baseline symptom severity was associated with slight increases in last reported anxiety and depression. Previous research suggests greater improvements for users with elevated baseline anxiety and depression symptoms (^44,45^) although findings are mixed (^7,42^). Compared to the structured computerized interventions evaluated in e-mental health research (^44,45^), Togetherall is designed as a supportive platform and not a modular intervention, nonetheless, Hensel et al. (^56^) reported decreases in anxiety and depression scores at 3-months follow-up after Togetherall use. Most members did not complete more than one clinical test in their service journey, limiting inferences that can be drawn regarding symptom score changes. It is possible that Togetherall members chose to take clinical tests only during moments of distress. Further, the study was a naturalistic design, without standardized measurement intervals. Members who experienced benefit and improvement may therefore have been systematically less likely to take a second test to evidence improvement. It is also possible that a subgroup of people with continued distress also took additional clinical tests to increase self-awareness. More controlled outcome collection at defined times could improve the accuracy of effectiveness estimates.

We note number of self-help materials accessed predicted decreased final anxiety scores. These results support evidence that adherence to e-mental health programmes predicts greater symptom improvements in young people (^16,35–37^). Thus, our findings suggest engagement with Togetherall self-help components improves anxiety outcomes. In contrast, engagement did not predict decreased depression scores, contrary to previous evidence from digital interventions (^37,79^ but see: ^83^). However, final depression scores were significantly lower for users who accessed more self-help materials in complete-case analyses. This observation suggests engagement with self-help on the platform could relate to reduction in depressive symptoms. However, results should be interpreted cautiously due to the different composition of primary and influence analyses. The level of participation with Togetherall components might be the active mechanisms of change, rather than simply time spent on the platform. Other factors may confound results, such as screen time exposure, affected sleep or various concurrent online activity (^17^). Studying content and number of posts in Togetherall forums, or recording alternative measures of engagement could clarify these questions.

Differing predictors of anxiety and depression outcomes also highlight the importance of tailoring approaches on generic platforms to different presentations. Research can be used to guide and inform the development and improvement of digital services over time. An important finding of this study is the evidence for real life engagement: online; anonymously; among a group of young people whose opportunity to share what they are going through and receive support might otherwise be absent. The current study suggests widespread engagement among a digital mental health community in this subset of young people.

Our sample comprised users referred from UK CAMH-services, rather than most existing e-mental health data from healthy non-referred adolescents (^3^). Further, the study addressed a gap in the literature around naturalistic engagement with e-mental health services (^33^). We controlled for different durations of Togetherall membership, accounting for their variability and effects on symptom change. Moreover, to control for biases in complete-case analyses (^84^) we also modelled imputed baseline data, offering a more precise method of missing data handling(^46,84,85^), suitable for observational studies (^86^). As with other internet interventions, our data was skewed and with high variability in usage metrics (^33,59,70^). Future studies could implement mixed effects models to estimate within-participant variability and missing data (^46,70^).

Due to the cross-sectional design we cannot infer causal relationships on Togetherall’s effectiveness. Furthermore, the clinical significance of the predicted changes in anxiety and depression is ambiguous considering reliable change indices of 4 points (GAD-7) for anxiety and 6 points (PHQ-9) for depression (^87^). It is unclear whether individuals in our sample had diagnoses of anxiety and/or depression or whether symptoms were comorbid with other diagnoses. We also only assessed multiple anxiety and depression measures in a minority of participants, thus measurement bias was present. In future, to better track progress, engagement measures could be analyzed prior to completion of each questionnaire on the platform whilst controlling for the time lapse between completed questionnaires (^70^). We were also unable to control for members accessing additional Togetherall components, such as “Live Therapy”, forum posts, self-expression through digital art or percentage of guided course completion. These components may differentially capture engagement and could be explored in future research.

The multicomponent structure of Togetherall poses challenges to operationalizing optimal time of use, making it difficult to define ‘good’ engagement. Skewed measures of usage and the large proportion of users who did not access courses or self-help materials mirror inconsistent engagement patterns reported in e-mental health literature (^16,34,36^). Adherence to e-mental health in naturalistic environments is expected to be even lower than in controlled trials (^33^). However, for Togetherall, only 33.5% of our included sample discontinued use after first login - notably less than reported drop-out rates in a number of previous controlled studies of other digital interventions (^35,88,56^). Our results indicate Togetherall is accessible and acceptable to young people. Behaviour on the platform can predict symptom change, adding to the literature on the importance of studying engagement with e-mental health (^33^). Accessing Togetherall self-help materials predicted decreases in anxiety symptoms. Active engagement with Togetherall components might be more relevant to improved symptom outcomes. However, many adolescents in our sample did not access materials and drop-out rates from guided courses should still be examined. Our findings highlight that engagement with Togetherall materials can facilitate improved wellbeing, whilst also addressing the effects of additional platform features and confounding factors of online behaviour.

This echoes survey data demonstrating students with anxiety symptoms were more likely to use self-help resources whilst those with higher depression symptoms were more likely to access anonymous online chat (^89^). Additionally, adolescents who reported their gender as other showed elevated baseline depression, comparably to females and greater than males. This is consistent with survey results of higher depression and anxiety related to prior use of e-mental health services in students reporting “other” gender (^89^). Overall, our findings suggest Togetherall may offer a supportive community to address the mental health needs in gender-diverse population and calls for research exploring preferred Togetherall features in male and gender-diverse adolescents.

Future studies are needed to establish Togetherall effectiveness, adherence and acceptability in young people using robust RCTs with active comparison groups. Time periods for engagement with Togetherall could be defined (e.g. 3 and 6 months) allowing for multilevel analysis (^56,88^). Three months is half of the Togetherall license and parallel treatment time for many effective psychological interventions (^88,6,57^). Additional factors that might influence engagement with Togetherall and subsequent symptom improvements could also be examined, such as geographical location, guidance and peer support on the platform. Further research could assess outstanding Togetherall components or use alternative operationalizations of engagement. Qualitative methods could also explore motivations for and barriers to engagement with the platform.

Beyond examining individual predictors, embedding theoretical models in the study of engagement and outcome may reveal how interactions with digital platforms generate behavior change, such as Self-determination theory (^78,90,91^). Future studies could use this as a framework linked to Togetherall characteristics to facilitate optimal engagement and its relationship with improved wellbeing.

## Data Availability

The data reported in the study is held by Togetherall and was subject to a data sharing agreement with the University of Edinburgh. The data remains with Togetherall.

## Supplemental Material

**Supplemental Figure 1:**
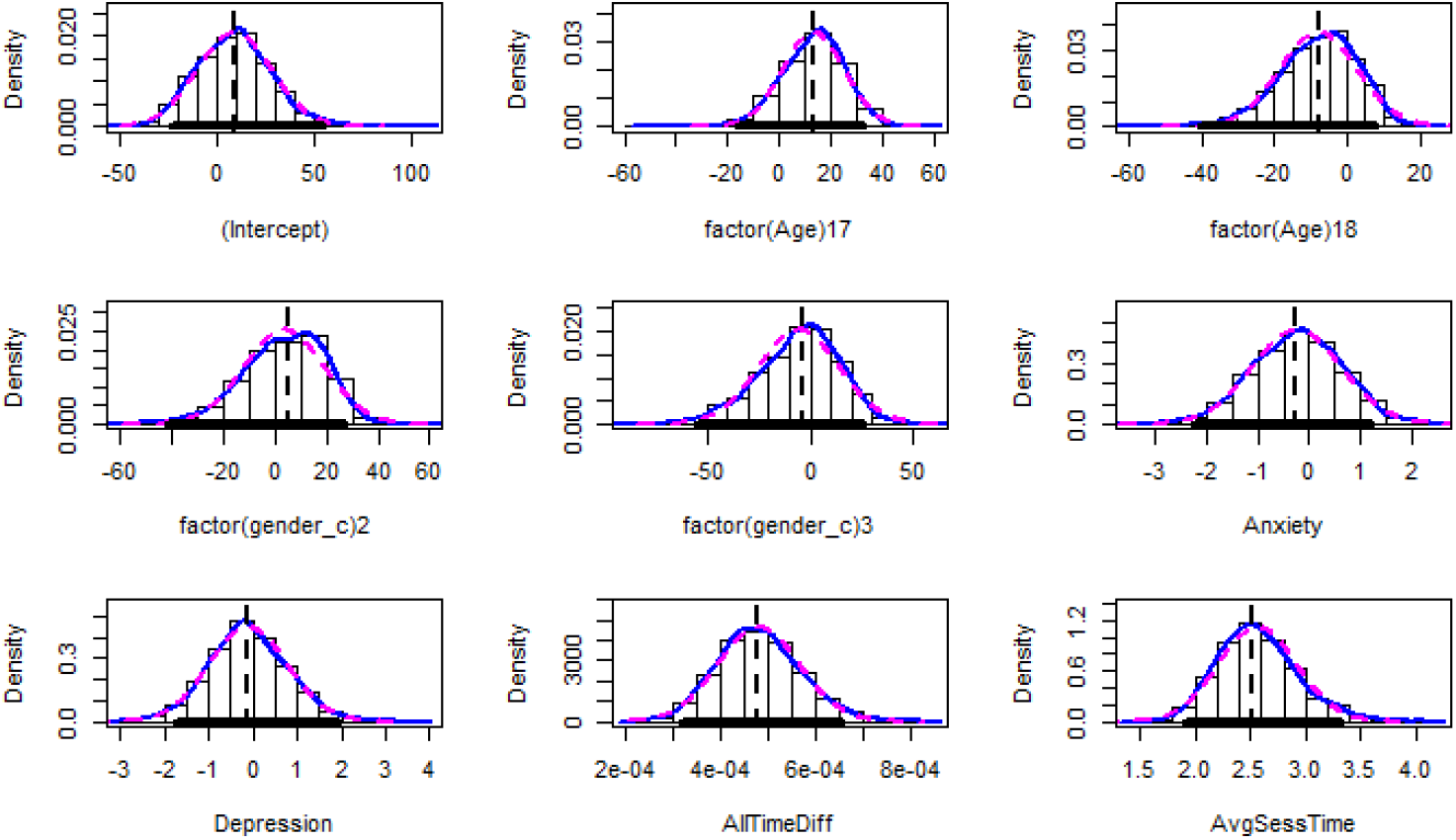
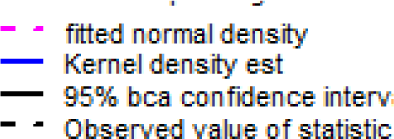
Kernel density plots for bootstrapped and observed distribution

**Supplemental Figure 2:**
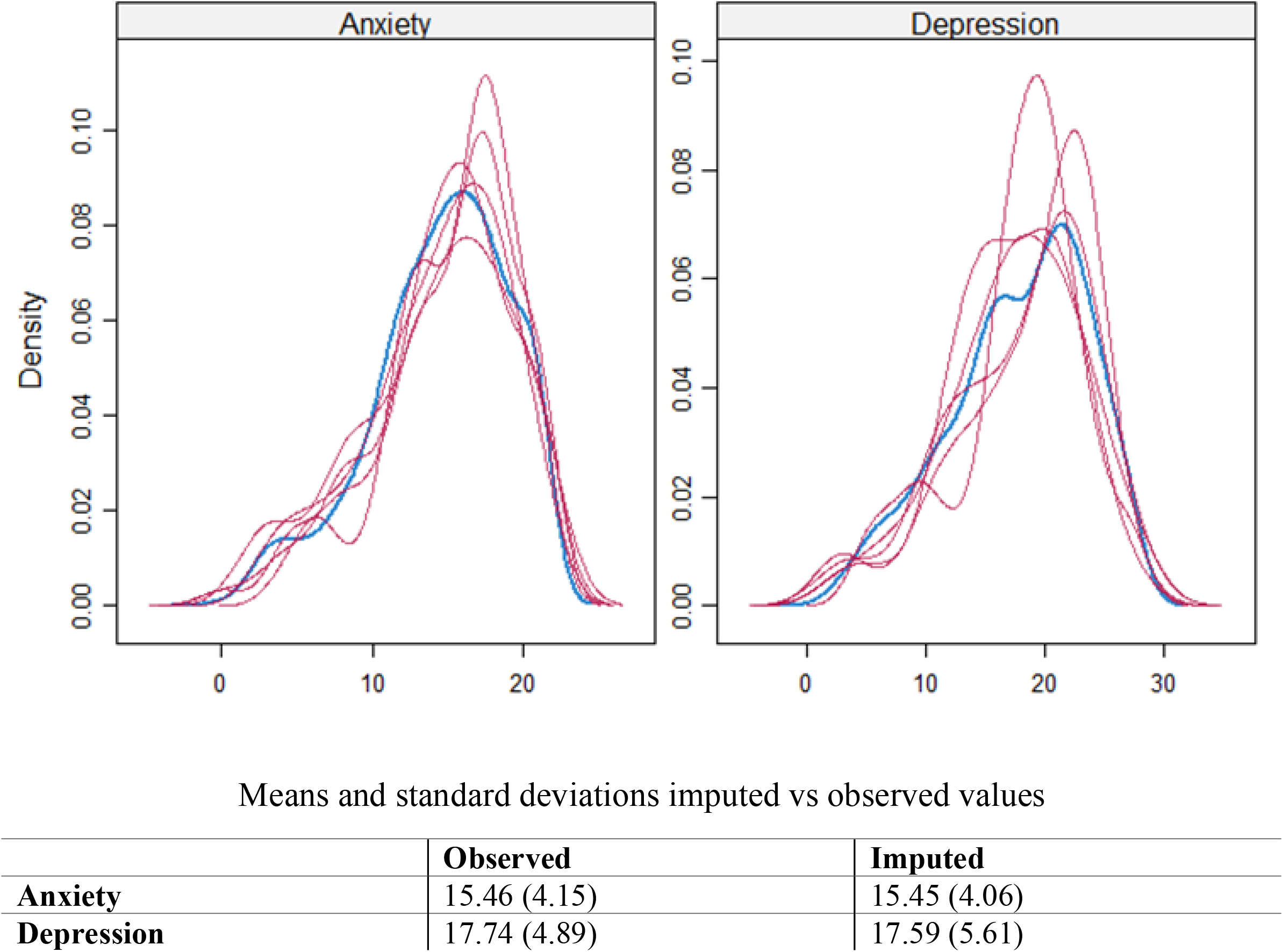
Distribution of observed (blue) vs imputed data (magenta)

**Supplemental Table 1.**
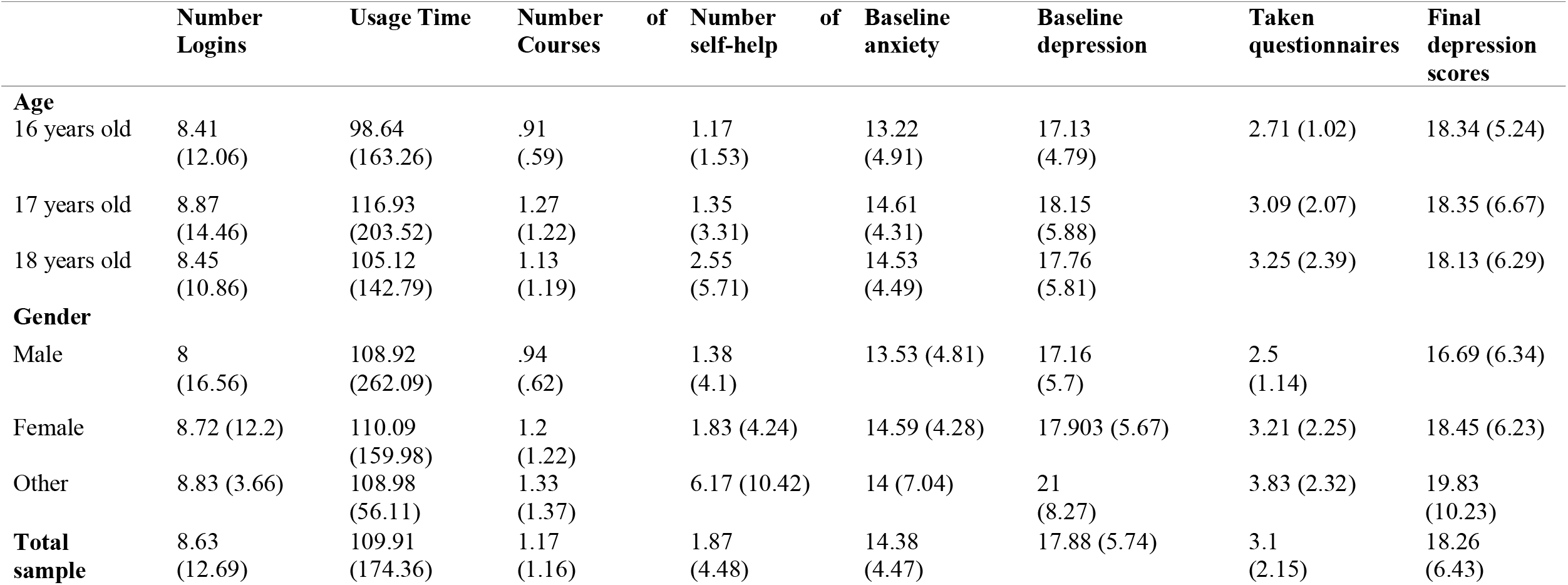
Mean and standard deviations of usage metrics and depression outcomes (n=245)

**Supplemental Table 2.**
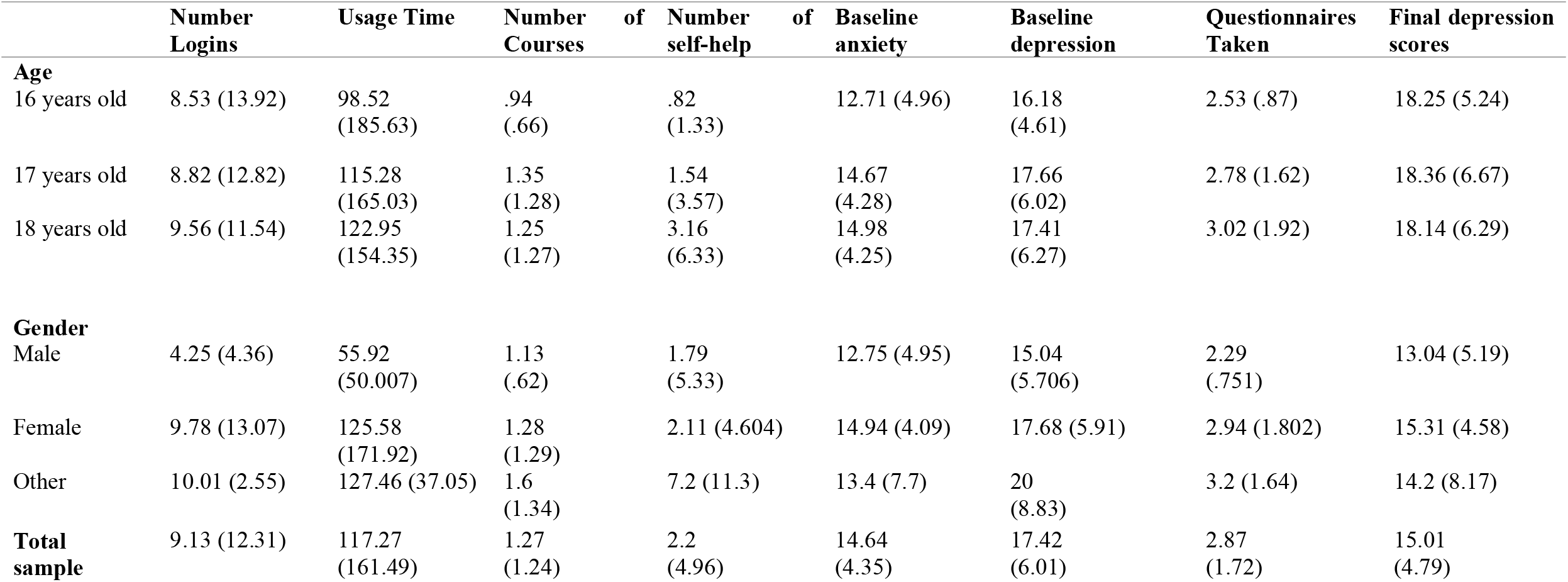
Means and standard deviations of usage metrics and anxiety outcomes (N=200)

|  | Number Logins | Usage Time | Number of Courses | Number of self-help | Baseline anxiety | Baseline depression | Questionnaires Taken | Final depression scores |
| --- | --- | --- | --- | --- | --- | --- | --- | --- |
| **Age** |  |  |  |  |  |  |  |  |
| 16 years old | 8.53 (13.92) | 98.52 (185.63) | .94 (.66) | .82 (1.33) | 12.71 (4.96) | 16.18 (4.61) | 2.53 (.87) | 18.25 (5.24) |
| 17 years old | 8.82 (12.82) | 115.28 (165.03) | 1.35 (1.28) | 1.54 (3.57) | 14.67 (4.28) | 17.66 (6.02) | 2.78 (1.62) | 18.36 (6.67) |
| 18 years old | 9.56 (11.54) | 122.95 (154.35) | 1.25 (1.27) | 3.16 (6.33) | 14.98 (4.25) | 17.41 (6.27) | 3.02 (1.92) | 18.14 (6.29) |
| **Gender** |  |  |  |  |  |  |  |  |
| Male | 4.25 (4.36) | 55.92 (50.007) | 1.13 (.62) | 1.79 (5.33) | 12.75 (4.95) | 15.04 (5.706) | 2.29 (.751) | 13.04 (5.19) |
| Female | 9.78 (13.07) | 125.58 (171.92) | 1.28 (1.29) | 2.11 (4.604) | 14.94 (4.09) | 17.68 (5.91) | 2.94 (1.802) | 15.31 (4.58) |
| Other | 10.01 (2.55) | 127.46 (37.05) | 1.6 (1.34) | 7.2 (11.3) | 13.4 (7.7) | 20 (8.83) | 3.2 (1.64) | 14.2 (8.17) |
| **Total sample** | 9.13 (12.31) | 117.27 (161.49) | 1.27 (1.24) | 2.2 (4.96) | 14.64 (4.35) | 17.42 (6.01) | 2.87 (1.72) | 15.01 (4.79) |

**Supplemental Table 3:** Robust regression coefficient for predicted usage time, with no outliers (n=602)

|  | Average bootstrap estimate | Bootstrap standard error | Asymptotic standard error | Bootstrap confidence intervals (95%) | Percentile intervals (95%) | P-values |
| --- | --- | --- | --- | --- | --- | --- |
| **Constant** | 15.21 | 17.17 | 21.4 | [-13.25;43.23] | [-11.52;45.58] | .51 |
| **Age (Reference category = 16 years old)** |  |  |  |  |  |  |
| 17 years old | 12.33 | 11.7 | 12.72 | [-6.38; 32.11] | [-8.16; 30.76] | .34 |
| 18 years old | -7.16 | 10.46 | 12.76 | [-24.1; 10.32] | [-25.76;7.66] | .57 |
| **Gender (Reference category = Male)** |  |  |  |  |  |  |
| Female | 5.69 | 15.27 | 10.83 | [-18.86; 31.36] | [-22.05;27.21] | .61 |
| Other | -1.59 | 19.52 | 24.89 | [-33.21; 31.01] | [-37.2; 27.42] | .95 |
| **Baseline depression** | -.425 | -.826 | .916 | [-2.17, .86] | [-1.81; .95] | .64 |
| **Baseline anxiety** | -.397 | .772 | 1.14 | [-1.86,1.57] | [-1.59; .94] | .79 |
| **Total time registered** | .001 | <0.001 | <0.001 | [<.001, .001] | [<.001; .001] | <.001*** |
| **Average session duration** | 2.409 | .342 | .45 | [1.83, 3.11] | [1.92; 3.01] | <.001*** |

P-value significance: \*\*\*<.001; \*\*<.01; \*<.05

**Supplemental Table 4:**
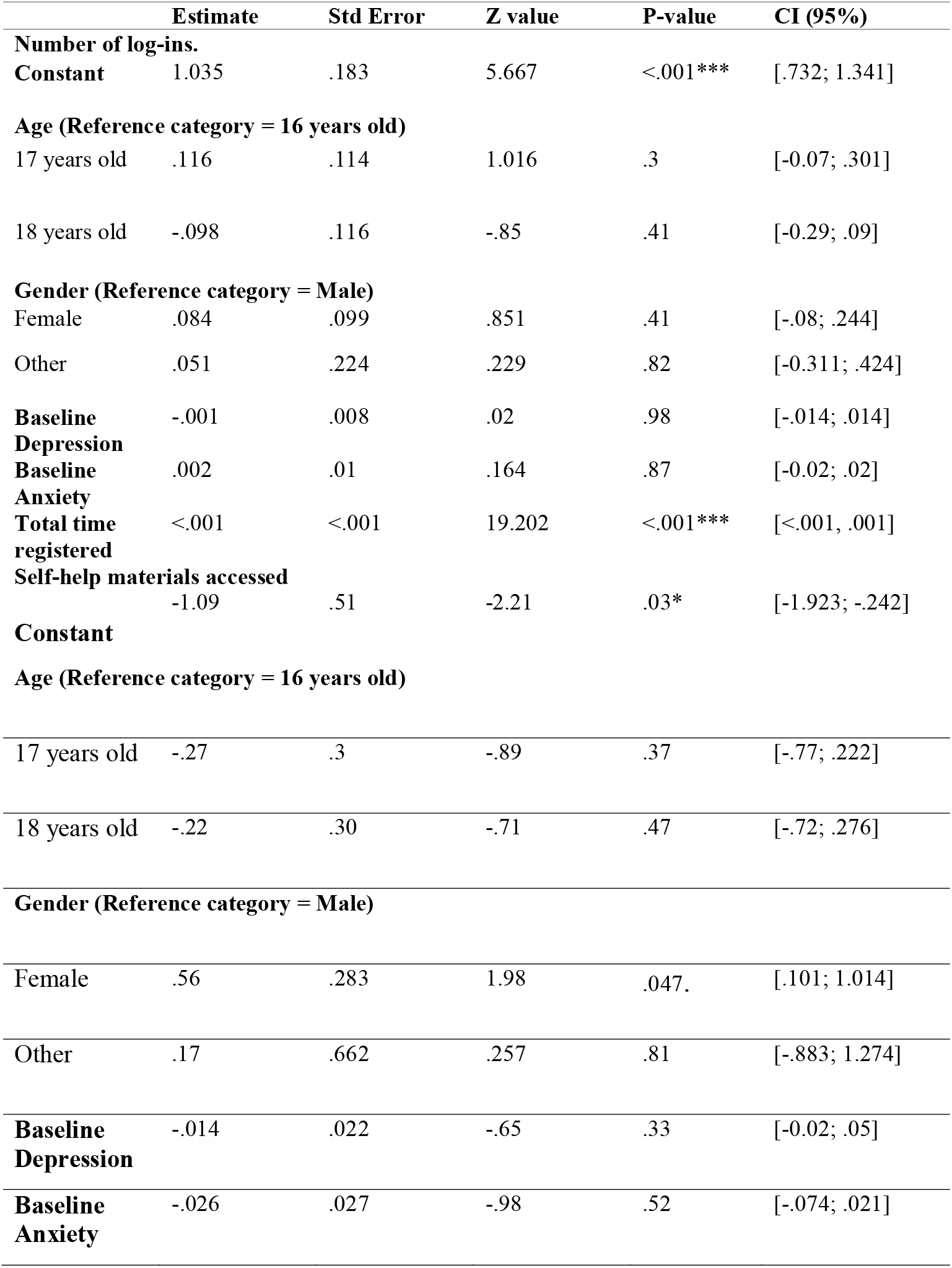
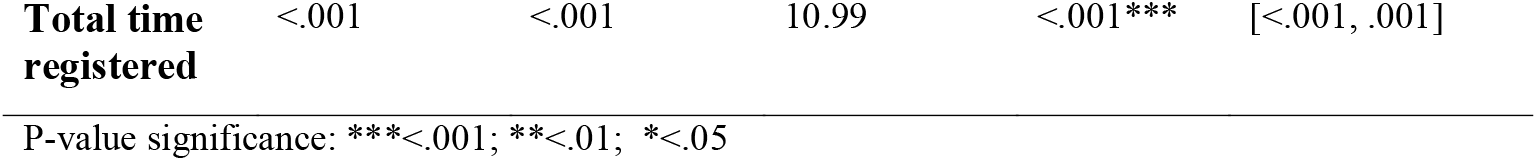
Negative binomial regression predicting number of logins, and self-help materials accessed, no outliers (n=602)

**Supplemental Table 5.** Stepwise regression coefficients for predictors of final depression (n=239) and anxiety scores (n=194), no outliers

|  | Estimate | Std Error | T value | P-value | CI (95 %) |
| --- | --- | --- | --- | --- | --- |
| **Final Depression Scores** |  |  |  |  |  |
| **Constant** | .39 | 1.215 | .321 | .75 | [-2.396; 1.62] |
| **Gender (Reference category = Male)** |  |  |  |  |  |
| Female | 1.132 | .737 | 1.536 | .13 | [-.085; 2.349] |
| Other | .89 | 1.724 | .52 | .61 | [-1.96; 3.74] |
| **Baseline Anxiety** | .126 | .066 | 1.909 | .06. | [.017; .235] |
| **Baseline Depression** | .772 | .052 | 15.883 | <.001*** | [.74; .912] |
| **Usage Time** | .004 | .002 | 2.328 | .02* | [.001; .007] |
| **Average Session Duration** | .069 | .028 | 2.482 | .02* | [.023, .114] |
| **Number of accessed self-help** | -.171 | .077 | -2.239 | .03* | [-0.298, -.045] |
| **Number of accessed courses** | -.02 | .265 | -.063 | .95 | [-.454; .421] |
| **Final Anxiety score** |  |  |  |  |  |
| **Constant** | 2.64 | .815 | 3.221 | .001** | [1.307, 3.982] |
| **Baseline Anxiety** | 0.646 | .056 | 11.56 | <.001*** | [.554, .74] |
| **Baseline Depression** | 0.182 | .04 | 4.58 | <.001*** | [.117, .249] |
| **Number of accessed self-help** | -.093 | .047 | -1.99 | 0.04* | [-.17, -.016] |

P-value significance: \*\*\*<.001; \*\*<.01; \*<.05. *Depression model: R2 =.66, Residual Std Error=3.75, DF=230; Anxiety Model: R2 =.62, Residual Std Error=2.81, DF=190*

**Supplemental Table 6:** Robust regression coefficients for predicted usage time, complete-case analyses (n=483)

|  | **Average**<br>**bootstrap**<br>**estimate** | **Bootstrap**<br>**standard**<br>**error** | **Asympt**<br>**otic**<br>**standar**<br>**d error** | **Bootstrap**<br>**confidence**<br>**intervals**<br>**(95%)** | **Percentile**<br>**intervals**<br>**(95%)** | **P-**<br>**values** |
| --- | --- | --- | --- | --- | --- | --- |
| **Constant** | 26.5 | 23.95 | 22.8 | [-13.34, 65.45] | [-12.06,68.77] | .24 |
| **Age (Reference category = 16 years old)** |  |  |  |  |  |  |
| 17 years old | 6.18 | 14.21 | 13.83 | [-17.25,29.45] | [-19.21,27.95] | .66 |
| 18 years old | -9.43 | 13.39 | 14.08 | [-31.36,12.72] | [-33.06,10.43] | .49 |
| **Gender (reference category = male)** |  |  |  |  |  |  |
| Female | 27.95 | 9.05 | 11.93 | [13.45,43.23] | [13.29, 43.37] | .02* |
| Other | 26.27 | 15.25 | 27.38 | [1.39,51.54] | [.99, 51.53] | .34 |
| **Baseline**<br>**depression** | -.69 | .91 | .974 | [-2.15, .79] | [-2.19, .753] | .45 |
| **Baseline**<br>**anxiety** | -.27 | 1.04 | 1.21 | [-1.99, 1.44] | [-1.98, 1.37] | .82 |
| **Total**<br>**time registered** | <0.001 | <0.001 | <0.001 | [<.001, .001] | [<.001, .001] | <.001**<br>* |
| **Average session**<br>**time** | 2.49 | .389 | .47 | [1.83, 3.11] | [1.92, 3.20] | <.001<br>*** |

$R^2 = .34$ [.16;.47], *Residual Std Error*=.08. P-value significance: \*\*\*<.001; \*\*<.01; \*<.05.

**Supplemental Table 7:**
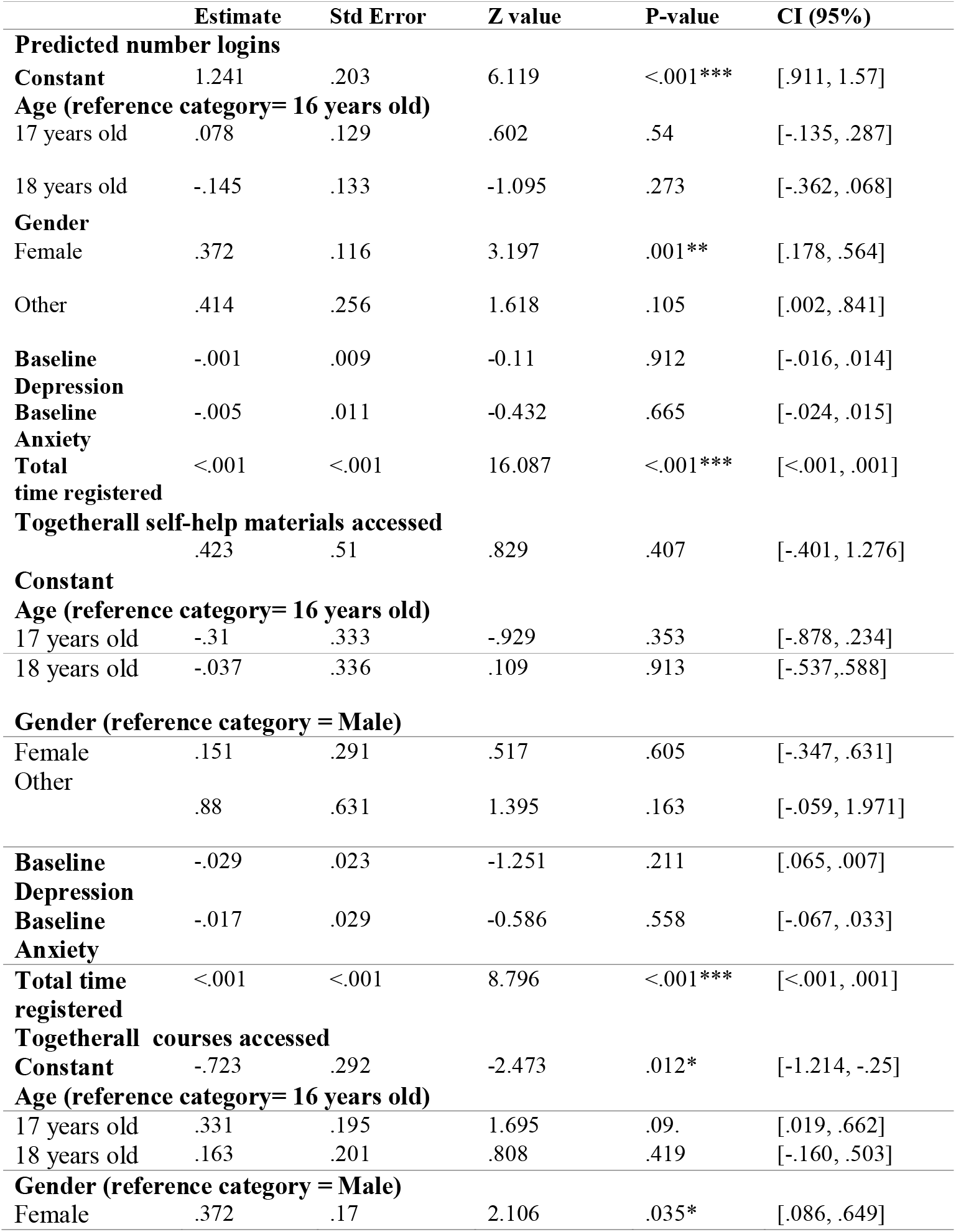
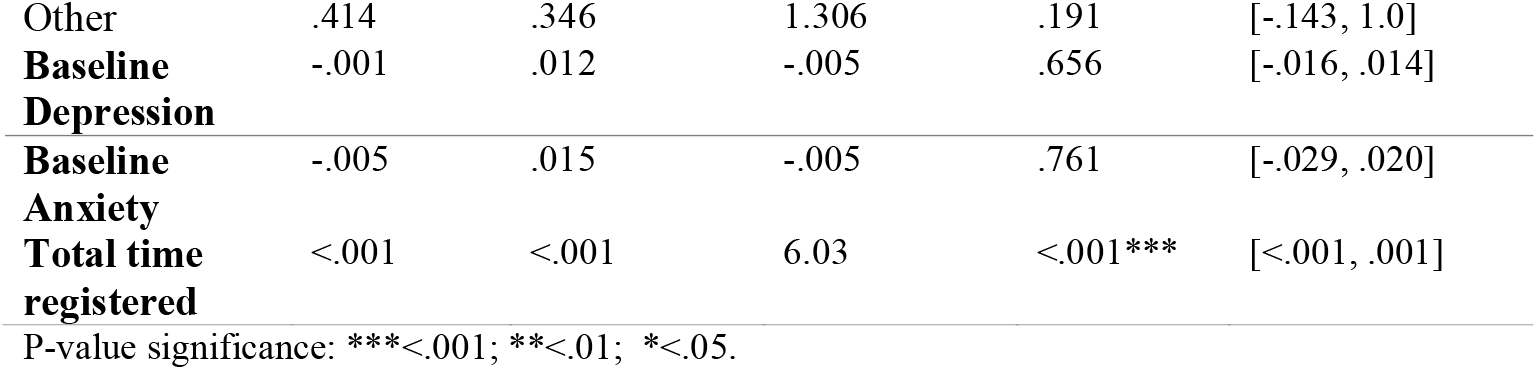
Negative binomial regression coefficients for predicted number logins, Togetherall activities accessed and Togetherall courses accessed, (Complete-case Analyses; n=483)

**Supplemental Table 8.** Stepwise regression coefficients for predictors of final depression (n=235) and anxiety (n=196) symptom scores, Completer analysis.

|  | **Estimate** | **Std Error** | **T value** | **P-value** | **CI (95 %)** |
| --- | --- | --- | --- | --- | --- |
| **Final depression scores** |  |  |  |  |  |
| **Constant** | .929 | 1.135 | .818 | .414 | [-1.308, 3.166] |
| **Baseline Anxiety** | .191 | .07 | 2.722 | .007** | [.053, .329] |
| **Baseline Depression** | .776 | .055 | 14.227 | <.001*** | [.669, .884] |
| **Usage Time** | .006 | .002 | 2.855 | .005** | [.002, .009] |
| **Average Session Duration** | .055 | .029 | 1.896 | .059. | [-.002, .113] |
| **Number of accessed self-help** | -.117 | .067 | -1.747 | .082. | [-.25, .015] |
| **Number of accessed courses** | -.359 | .251 | -1.429 | .154 | [-.854, .136] |
| **Final Anxiety Scores** |  |  |  |  |  |
| **Constant** | 2.986 | .898 | 3.326 | .001** | [1.215, 4.757] |
| **Baseline Anxiety** | .665 | .063 | 10.538 | <.001*** | [.541, .790] |
| **Baseline Depression** | .144 | .046 | 3.166 | .002** | [.054, .234] |
| **Number of accessed self-help** | -.118 | .047 | -2.512 | .013* | [-.211, -.025] |

*Depression score analyses: $R^2 = .62$ , Residual Std Error=4.02, DF=228. Anxiety score analyses: $R^2 = .56$ , Residual Std Error=3.232, DF=192. P-value significance: \*\*\*<.001; \*\*<.01; \*<.05.*

## Notes

### Competing Interest Statement

The authors have declared no competing interest.

### Funding Statement

The project was completed as part of the first author's (Nushka Marinova) MSc studies. The training programme was funded by NHS Education Scotland.

### Author Declarations

University of Edinburgh School of Health in Social Science Ethics Committee.

## References

1. Erskine HE, Moffitt TE, Copeland WE, et al. A heavy burden on young minds: the global burden of mental and substance use disorders in children and youth. Psychol Med. 2018;45(7):1551–1563. doi:10.1017/S0033291714002888.A

2. Polanczyk G V, Salum GA, Sugaya LS, Caye A, Rohde LA. Annual Research Review□: A meta-analysis of the worldwide prevalence of mental disorders in children and adolescents. J Child Psychol Psychiatry. 2015;56(3):345–365. doi:10.1111/jcpp.12381

3. Grist R, Porter J, Stallard P. Mental Health Mobile Apps for Preadolescents and Adolescents□: A Systematic Review Corresponding Author□: J Med Internet Res. 2017;19(5):e176. doi:10.2196/jmir.7332

4. Mclaughlin KA, King K. Developmental Trajectories of Anxiety and Depression in Early Adolescence. J Abnorm Child Psychol. 2015;43:311–323. doi:10.1007/s10802-014-9898-1

5. Gladstone TRG, Connor EEO, Beardslee WR. T h e Prev e n t i o n of Adolescent D e p re s s i o n. Psychiatr Clin NA. 2011;34(1):35–52. doi:10.1016/j.psc.2010.11.015

6. Cox G, Callahan P, Churchill R, et al. Psychological therapies versus antidepressant medication, alone and in combination for depression in children and adolescents (Review). Cochrane Libr. 2014. doi:10.1002/14651858.CD008324.pub3. www.cochranelibrary.com

7. Vigerland S, Lenhard F, Bonnert M, et al. Internet-delivered cognitive behavior therapy for children and adolescents□: A systematic review and meta-analysis. Clin Psychol Rev. 2016;50:1–10. doi:10.1016/j.cpr.2016.09.005

8. Brownlie E, Beitchman JH, Chaim G, Wolfe DA, Rush B, Henderson J. Early Adolescent Substance Use and Mental Health Problems and Service Utilisation in a School-based Sample. Can J Psychiatry. 2019;64(2):116–125. doi:10.1177/0706743718784935

9. Costello EJ, Ph D, He J, et al. Services for Adolescents With Psychiatric Disorders□: 12-Month Data From the National Comorbidity Survey – Adolescent. Psychiatr Serv. 2014;65(3):359–366. doi:10.1176/appi.ps.201100518

10. England El, Mughal F. Underprovision of mental health services for children and young people. Br J Gen Pract. 2019:112–113. doi:https://doi.org/10.3399/bjgp19X701381

11. Rocha TB, Graeff-martins AS. Provision of mental healthcare for children and adolescents□: a worldwide view. Curr Opin Psychiatry. 2015;28(4):330–335. doi:10.1097/YCO.0000000000000169

12. Fonagy P, Pugh K, Herlihy AO. The Children and Young People ‘ s Improving Access to Psychological Therapies (CYP IAPT) Programme in England. In: Skuse D, Bruce H, Dowdney L, eds. Child Psychology and Psychiatry: Frameworks for Clinical Training and Practice. John Wiley & Sons, Ltd; 2017:429–435.

13. Gulliver A, Griffiths KM, Christensen H. Perceived barriers and facilitators to mental health help-seeking in young people□: a systematic review. BMC Psychiatry. 2010;113(10):1–9. doi:doi:10.1186/1471-244X-10-113

14. Sheppard R, Deane FP, Ciarrochi J. Unmet need for professional mental health care among adolescents with high psychological distress. Aust New Zeal J Psychiatry. 2018;52(1):59–67. doi:10.1177/0004867417707818

15. Macdonell KW, Prinz RJ. A Review of Technology-Based Youth and Family-Focused Interventions. Clin Child Fam Psychol Rev. 2018;20(2):185–200. doi:10.1007/s10567-016-0218-x.A

16. Clarke AM, Kuosmanen T, Barry MM. A Systematic Review of Online Youth Mental Health Promotion and Prevention Interventions. Youth Adolesc. 2015;44:90–113. doi:10.1007/s10964-014-0165-0

17. Naslund JA, Gonsalves PP, Gruebner O, et al. Digital Innovations for Global Mental Health□: Opportunities for Data Science, Task Sharing, and Early Intervention. Curr Treat Options Psychiatry OnlineFIist. 2019:1–15. doi:10.1007/s40501-019-00186-8

18. Anderson M, Jiang J. Teens, Social Media & Technology 2018. https://www.pewinternet.org/2018/05/31/teens-social-media-technology-2018/. Published 2018. Accessed November 11, 2019.

19. Hoare E, Milton K, Foster C, Allender S. Depression, psychological distress and Internet use among community-based Australian adolescents: A cross-sectional study. BMC Public Health. 2017;17(1):1–10. doi:10.1186/s12889-017-4272-1

20. Ridout B, Campbell A. The Use of Social Networking Sites in Mental Health Interventions for Young People□: Systematic Review Corresponding Author□: J Med Internet Res. 2018;20:1–11. doi:10.2196/12244

21. Wozney L, Mcgrath PJ, Gehring ND, Bennett K, Huguet A, Newton AS. eMental Healthcare Technologies for Anxiety and Depression in Childhood and Adolescence□: Systematic Review of Studies Reporting Implementation Outcomes Corresponding Author□: JMIR Ment Heal. 2018;5(2):248. doi:10.2196/mental.9655

22. Välimäki M, Katriina A, Minna A, Mari L. Web-Based Interventions Supporting Adolescents and Young People With Depressive Symptoms□: Systematic Review and Corresponding Author□: JMIR MHEALTH UHEALTH. 2017;5(12). doi:10.2196/mhealth.8624

23. Olthuis J, Watt M, Bailey K, Hayden J, Stewart S. Therapist-supported Internet cognitive behavioural therapy for anxiety disorders in adults (Review). Cochrane Libr. 2016. doi:10.1002/14651858.CD011565.pub2. www.cochranelibrary.com

24. Dea BO, Calear AL, Perry Y. Is e-health the answer to gaps in adolescent mental health service provision□? Curr Opin Psychiatry. 2015;28(4):336–342. doi:10.1097/YCO.0000000000000170

25. Domhardt M, Steubl L, Baumeister H. Internet-and Mobile-Based Interventions for Mental and Somatic Conditions in Children and Adolescents A Systematic Review of Meta-analyses. Z Kinder Jugendpsychiatr Psychother. 2018:1–14. doi:10.1024/1422-4917/a000625

26. Ebert DD, Zarski A, Christensen H. Internet and Computer-Based Cognitive Behavioral Therapy for Anxiety and Depression in Youth□: A Meta-Analysis of Randomized Controlled Outcome Trials. PLoS One. 2015;10(3):1–15. doi:10.1371/journal.pone.0119895

27. Hollis C, Falconer CJ, Martin JL, et al. Annual Research Review□: Digital health interventions for children and young people with mental health problems – a systematic and meta-review. J Child Psychol Psychiatry. 2017;58(4):474–503. doi:10.1111/jcpp.12663

28. Carlbring P, Andersson G, Cuijpers P, Riper H. Internet-based vs. face-to-face cognitive behavior therapy for psychiatric and somatic disorders□: an updated systematic review and meta-analysis. Cogn Behav Ther. 2018;6073:1–21. doi:10.1080/16506073.2017.1401115

29. Ebert DD, Daele T Van, Nordgreen T, et al. Internet-and Mobile-Based Psychological Interventions□: Applications, Efficacy, and Potential for Improving Mental Health A Report of the EFPA E-Health Taskforce. Eur Psychol. 2018;23(2):167–187. https://doi.org/10.1027/1016-9040/a000318.

30. Owens VAM, Hadjistavropoulos HD, Schneider LH, et al. Transdiagnostic, internet-delivered cognitive behavior therapy for depression and anxiety□: Exploring impact on health anxiety. Internet Interv. 2019;15(January):60–66. doi:10.1016/j.invent.2019.01.001

31. Weisel KK, Zarski A, Berger T, et al. E ffi cacy and cost-e ff ectiveness of guided and unguided internet-and mobile-based indicated transdiagnostic prevention of depression and anxiety (ICare Prevent): A three-armed randomized controlled trial in four European countries. Internet Interv. 2019;16(April 2018)52–64. doi:10.1016/j.invent.2018.04.002

32. Heber E, Ebert DD, Lehr D, Cuijpers P, Berking M. The Benefit of Web-and Computer-Based Interventions for Stress□: A Systematic Review and Meta-Analysis. J Med Internet Res. 2017;19(2):e32. doi:10.2196/jmir.5774

33. Fleming T, Bavin L, Lucassen M, et al. Beyond the Trial□: Systematic Review of Real-World Uptake and Engagement With Digital Self-Help Interventions for Depression, Low Mood, or Anxiety. J Med Internet Res. 2018;20(6):e199. doi:10.2196/jmir.9275

34. Sieverink F, Kelders SM, Gemert-pijnen JEWC Van. Clarifying the Concept of Adherence to eHealth Technology□: Systematic Review on When Usage Becomes Adherence. J Med Internet Res. 2017;19(12):e402. doi:10.2196/jmir.8578

35. Neil AL, Batterham P, Christensen H, Bennett K, Griffiths KM, Neil AL. Predictors of Adherence by Adolescents to a Cognitive Behavior Therapy Website in School and Community-Based Settings Corresponding Author□: J Med Internet Res. 2009;11(1):e6. doi:10.2196/jmir.1050

36. Donkin L, Christensen H, Naismith SL, et al. A Systematic Review of the Impact of Adherence on the Effectiveness of e-Therapies. J Med Internet Res. 2011;13(3):e52. doi:10.2196/jmir.1772

37. Calear AL, Christensen H, Mackinnon A, Griffiths KM. Adherence to the MoodGYM program□: Outcomes and predictors for an adolescent school-based population. J Affect Disord. 2013;147:338–344. doi:10.1016/j.jad.2012.11.036

38. Karyotaki E, Riper H, Twisk J, et al. Efficacy of Self-guided Internet-Based Cognitive Behavioral Therapy in the Treatment of Depressive Symptoms A Meta-analysis of Individual Participant Data. JAMA Psychiatry. 2017;74(4):351–359. doi:10.1001/jamapsychiatry.2017.0044

39. Alaoui S El, Ljótsson B, Hedman E, Svanborg C, Kaldo V, Lindefors N. Predicting Outcome in Internet-Based Cognitive Behaviour Therapy for Major Depression□: A Large Cohort Study of Adult Patients in Routine Psychiatric Care. PLoS One. 2016;11(9):1–16. doi:10.1371/journal.pone.0161191

40. Sieverink F, Kelders S, Poel M, Gemert-pijnen L Van. Opening the Black Box of Electronic Health□: Collecting, Analyzing, and Interpreting Log Data. JMIR Res Protoc. 2017;6(8):e156. doi:10.2196/resprot.6452

41. Podina IR, Mogoase C. A Meta-Analysis on the Efficacy of Technology Mediated CBT for Anxious Children and Adolescents. J Ration Cogn Ther. 2016;34(1):31–50. doi:10.1007/s10942-015-0228-5

42. Pennant ME, Loucas CE, Whittington C, et al. Behaviour Research and Therapy Computerised therapies for anxiety and depression in children and young people□: A systematic review and meta-analysis. Behav Res Ther. 2015;67:1–18. doi:10.1016/j.brat.2015.01.009

43. Khan HA, Bernstein K. Online Therapy for Adolescent Mental Health. In:Moreno M, Radovic A, eds. Technology and Adolescent Mental Health. Vol 16. Springer, Cham; 2018:217–236. doi:10.1007/978-3-319-69638-6_16

44. Stjerneklar S, Hougaard E, Thastum M. Guided internet-based cognitive behavioral therapy for adolescent anxiety□: Predictors of treatment response. Internet Interv. 2019;15(January)116–125. doi:10.1016/j.invent.2019.01.003

45. Grist R, Croker A, Denne M, Stallard P. Technology Delivered Interventions for Depression and Anxiety in Children and Adolescents□: A Systematic Review and Meta-analysis. Clin Child Fam Psychol Rev. 2018;0(0):0. doi:10.1007/s10567-018-0271-8

46. Christensen H, Griffiths KM, Farrer L. Adherence in Internet Interventions for Anxiety and Depression□: Systematic Review. J Med Internet Res. 2009;11(2):e13. doi:10.2196/jmir.1194

47. Christie S. Big White Wall□: transforming mental health services through digital technologies. 2014. doi:10.1108/MHSI-07-2013-0024

48. Cheek C, Bridgman H, Fleming T, et al. Views of Young People in Rural Australia on SPARX, a Fantasy World Developed for New Zealand Youth With Depression Corresponding Author□: JMIR Serious Games. 2014;2(1):23. doi:10.2196/games.3183

49. Ho J, Corden ME, Caccamo L, et al. Design and evaluation of a peer network to support adherence to a web-based intervention for adolescents. Internet Interv. 2016;6:50–56. doi:10.1016/j.invent.2016.09.005

50. Harding C, Chung H. Behavioral health support and online peer communities□: international experiences. mHealth. 2016;2(43):1–6. doi:10.21037/mhealth.2016.10.04

51. Barton J, Henderson J. Peer Support and Youth Recovery: A Brief Review of the Theoretical Underpinnings and Evidence. Can J Fam Youth. 2016;8(1):1–17. doi:10.1037/a0031920

52. Rice SM, Clin M, Goodall J, et al. Online and Social Networking Interventions for the Treatment of Depression in Young People□: A Systematic Review Corresponding Author□: J Med Internet Res. 2014;16(9):e207. doi:10.2196/jmir.3304

53. Lawlor A, Kirakowski J. Computers in Human Behavior Online support groups for mental health□: A space for challenging self-stigma or a means of social avoidance□? Comput Human Behav. 2014;32:152–161. doi:10.1016/j.chb.2013.11.015

54. Easton K, Psych M, Diggle J, et al. Qualitative Exploration of the Potential for Adverse Events When Using an Online Peer Support Network for Mental Health□: Cross-Sectional Survey. JMIR Ment Heal. 2017;4(4):e49. doi:10.2196/mental.8168

55. Laurance BJ, Henderson S, Howitt PJ, et al. Patient Engagement: Four Case Studies That Highlight The Potential For Improved Health Outcomes And Reduced Costs. Health Aff. 2014;33(9):1627–1634. doi:10.1377/hlthaff.2014.0375

56. Hensel JM, Shaw J, Ivers NM, et al. A web-based mental health platform for individuals seeking specialized mental health care services: Multicenter pragmatic randomized controlled trial. J Med Internet Res. 2019;21(6):1–12. doi:10.2196/10838

57. Oud M, de Winter L, Vermeulen-Smit E, et al. Effectiveness of CBT for children and adolescents with depression: A systematic review and meta-regression analysis. Eur Psychiatry. 2019;57:33–45. doi:10.1016/j.eurpsy.2018.12.008

58. Wang Z, Whiteside S, Sim L, et al. Anxiety in Children. Rockville; 2017. doi:10.4324/9781315682471

59. Enrique Roig A, Palacios J, Ryan H, Richards D. The association between usage and outcomes of an online intervention for depression: how optimal dosage can help establish adherence (Preprint). J Med Internet Res. 2018;21. doi:10.2196/12775

60. Spitzer RL, Kroenke K, Williams JBW, Lo B. A Brief Measure for Assessing Generalized Anxiety Disorder. 2006;166:1092–1097.

61. Lowe B, Decker O, Muller S, et al. Validation and Standardization of the Generalized Anxiety Disorder Screener (GAD-7) in the General Population. Med Care. 2008;46(3):266–274. doi:10.1097/MLR.0b013e318160d093

62. Kroenke K, Spitzer RL, Williams JBW. The PHQ-9. 46202:606–613.

63. Kocalevent R, Ph D, H MP, et al. Standardization of the depression screener Patient Health Questionnaire (PHQ-9) in the general population. Gen Hosp Psychiatry. 2013;35(5):551–555. doi:10.1016/j.genhosppsych.2013.04.006

64. Imdadullah M, Aslam M, Altaf S. mctest: An R Package for Detection of Collinearity among Regressors. R J. 2016;8(2):499–509. https://journal.r-project.org/archive/2016/RJ-2016-062/index.html%0A.

65. Burgette LF, Reiter JP. Practice of Epidemiology Multiple Imputation for Missing Data via Sequential Regression Trees. Am J Epidemiol. 2010;172(9):1070–1076. doi:10.1093/aje/kwq260

66. Buuren van S, Groothuis-Oudshoorn K. Mice: Multivariate Imputation by Chained Equations in R. J Stat Softw. 2011;45(3):1–67. https://www.jstatsoft.org/v45/i03/.

67. Hartig F. DHARMa: Residual Diagnostics for Hierarchical (Multi-Level / Mixed) Regression Models. 2018. https://cran.r-project.org/package=DHARMa.

68. Vickers AJ, Altman DG. Analysing controlled trials with baseline and follow up measurements. BMJ. 2001;323:1123–1124.

69. Kuhn M. caret: Classification and Regression Training. 2019. https://cran.r-project.org/package=caret.

70. Carpenter J, Crutchley P, Zilca RD, et al. Seeing the “Big” Picture□: Big Data Methods for Exploring Relationships Between Usage, Language, and Outcome in Internet Intervention Data Corresponding Author□: Related Article□: J Med Internet Res. 2017;18(8):e241. doi:10.2196/jmir.5725

71. Su X, Tsai C. Outlier detection. 2011;1(June):261–268. doi:10.1002/widm.19

72. Cook RD. Detection of influential observation in linear regression. Technometrics. 1977;19(1):15–18.

73. Fox J, Weisberg S. Bootstrapping Regression Models in R (Web Appendix).; 2012. http://socserv.socsci.mcmaster.ca/jfox/Books/Companion/appendix.

74. Rickwood D, Webb M, Kennedy V, Telford N. Who Are the Young People Choosing Web-based Mental Health Support? Findings From the Implementation of Australia’s National Web-based Youth Mental Health Service, eheadspace. JMIR Ment Heal. 2016;3(3):e40. doi:10.2196/mental.5988

75. Do R, Park J, Lee S, Cho M, Kim J, Shin M. Adolescents ‘ Attitudes and Intentions toward Help-Seeking and Computer-Based Treatment for Depression. Psychiatry Investig. 2019;16(10):728–736. doi:https://doi.org/10.30773/pi.2019.07.17.4 Print

76. Thapar A, Collishaw S, Pine DS, Thapar AK. Depression in adolescence. Lancet. 2012;379(9820):1056–1067. doi:10.1016/S0140-6736(11)60871-4

77. Bennett SD, Cuijpers P, Ebert DD, et al. Practitioner Review: Unguided and guided self-help interventions for common mental health disorders in children and adolescents: a systematic review and meta-analysis. J Child Psychol Psychiatry Allied Discip. 2019;60(8):828–847. doi:10.1111/jcpp.13010

78. Farrer LM, Griffiths KM, Christensen H, Mackinnon AJ, Batterham PJ. Predictors of adherence and outcome in internet-based cognitive behavior therapy delivered in a telephone counseling setting. Cognit Ther Res. 2014;38(3):358–367. doi:10.1007/s10608-013-9589-1

79. Fuhr K, Schröder J, Berger T, et al. The association between adherence and outcome in an Internet intervention for depression. J Affect Disord. 2018;229(September 2017):443–449. doi:10.1016/j.jad.2017.12.028

80. Kleiboer A, Donker T, Seekles W, van Straten A, Riper H, Cuijpers P. A randomized controlled trial on the role of support in Internet-based problem solving therapy for depression and anxiety. Behav Res Ther. 2015;72:63–71. doi:10.1016/j.brat.2015.06.013

81. Radomski AD, Wozney L, McGrath P, et al. Design and delivery features that may improve the use of internet-based cognitive behavioral therapy for children and adolescents with anxiety: A realist synthesis with a persuasive systems design perspective. J Med Internet Res. 2019;21(2):1–21. doi:10.2196/11128

82. Kelders SM, Kok RN, Ossebaard HC, Van Gemert-Pijnen JEWC. Persuasive system design does matter: A systematic review of adherence to web-based interventions. J Med Internet Res. 2012;14(6). doi:10.2196/jmir.2104

83. Edmonds M, Hadjistavropoulos HD, Schneider LH, Dear BF, Titov N. Who benefits most from therapist-assisted internet-delivered cognitive behaviour therapy in clinical practice? Predictors of symptom change and dropout. J Anxiety Disord. 2018;54(July 2017):24–32. doi:10.1016/j.janxdis.2018.01.003

84. Vergouw D, Heymans MW, Van Der Windt DAWM, et al. Missing data and imputation: A practical illustration in a prognostic study on low back pain. J Manipulative Physiol Ther. 2012;35(6):464–471. doi:10.1016/j.jmpt.2012.07.002

85. Blankers M, Koeter MWJ, Schippers GM. Missing data approaches in eHealth research: Simulation study and a tutorial for nonmathematically inclined researchers. J Med Internet Res. 2010;12(5):1–13. doi:10.2196/jmir.1448

86. Kenward MG, Carpenter J. Multiple imputation: Current perspectives. Stat Methods Med Res. 2007;16(3):199–218. doi:10.1177/0962280206075304

87. National Collaborating Centre for Mental Health. The Improving Access to Psychological Therapies Manual: Appendices and Helpful Resources.; 2018. https://www.england.nhs.uk/wp-content/uploads/2018/06/iapt-manual-resources-v2.pdf.

88. Hensel JM, Shaw J, Ivers NM, et al. Extending access to a web-based mental health intervention□: who wants more, what happens to use over time, and is it helpful□? Results of a concealed, randomized controlled extension study. BMC Psychiatry. 2019:1–10. doi:https://doi.org/10.1186/s12888-019-2030-x

89. Toscos T, Coupe A, Flanagan M, Drouin M, Carpenter M. Teens Using Screens for Help□: Impact of Suicidal Ideation, Anxiety, and Depression Levels on Youth Preferences for Telemental Health Resources Corresponding Author□: JMIR Ment Heal. 2019;6(6):1–16. doi:10.2196/13230

90. Mohr DC, Burns MN, Schueller SM, Clarke G, Klinkman M. Behavioral Intervention Technologies: Evidence review and recommendations for future research in mental health. Gen Hosp Psychiatry. 2013;35(4):332–338. doi:10.1016/j.genhosppsych.2013.03.008

91. Pretorius C, Chambers D, Coyle D. Young people, Online Help-Seeking and Mental Health Difficulties: A Systematic Narrative Review (Preprint). J Med Internet Res. 2019;21. doi:10.2196/13873

